# Correlates of protection against SARS-CoV-2 Omicron variant and anti-spike antibody responses after a third/booster vaccination or breakthrough infection in the UK general population

**DOI:** 10.1101/2022.11.29.22282916

**Authors:** Jia Wei, Philippa C. Matthews, Nicole Stoesser, John N Newton, Ian Diamond, Ruth Studley, Nick Taylor, John I Bell, Jeremy Farrar, Jaison Kolenchery, Brian D. Marsden, Sarah Hoosdally, E Yvonne Jones, David I Stuart, Derrick W. Crook, Tim E. A. Peto, A. Sarah Walker, Koen B. Pouwels, David W. Eyre, the COVID-19 Infection Survey team

## Abstract

Following primary SARS-CoV-2 vaccination, understanding the relative extent of protection against SARS-CoV-2 infection from boosters or from breakthrough infections (i.e. infection in the context of previous vaccination) has important implications for vaccine policy. In this study, we investigated correlates of protection against Omicron BA.4/5 infections and anti-spike IgG antibody trajectories after a third/booster vaccination or breakthrough infection following second vaccination in 154,149 adults ≥18y from the United Kingdom general population. We found that higher anti-spike IgG antibody levels were associated with increased protection against Omicron BA.4/5 infection and that breakthrough infections were associated with higher levels of protection at any given antibody level than booster vaccinations. Breakthrough infections generated similar antibody levels to third/booster vaccinations, and the subsequent declines in antibody levels were similar to or slightly slower than those after third/booster vaccinations. Taken together our findings show that breakthrough infection provides longer lasting protection against further infections than booster vaccinations. For example, considering antibody levels associated with 67% protection against infection, a third/booster vaccination did not provide long-lasting protection, while a Delta/Omicron BA.1 breakthrough infection could provide 5-10 months of protection against Omicron BA.4/5 reinfection. 50-60% of the vaccinated UK population with a breakthrough infection would still be protected by the end of 2022, compared to <15% of the triple-vaccinated UK population without previous infection. Although there are societal impacts and risks to some individuals associated with ongoing transmission, breakthrough infection could be an efficient immune-boosting mechanism for subgroups of the population, including younger healthy adults, who have low risks of adverse consequences from infection.

## Introduction

Multiple SARS-CoV-2 vaccines have been developed and have been highly effective at reducing infections^1^ and associated hospitalisation and death^2–4^. However, waning of vaccine-induced immunity means optimal protection from vaccination may be relatively short-lived, with reduced effectiveness by 3-6 months reported after the second vaccination^5–7^ leading to widespread use of booster vaccinations. Reductions in vaccine effectiveness with time have been exacerbated by changes in circulating variants, with lower levels of protection against Delta compared to Alpha and further reductions against different Omicron variants^8^. However, alongside this, large numbers of “breakthrough” infections (i.e. natural infection in the context of previous vaccination) mean that an increasing proportion of the population has some existing immunity from earlier infections.

In those who have already received a primary vaccination course (typically two doses), understanding the relative extent of protection against further infection from booster vaccination has important implications for vaccine policy. One possible response to waning vaccine-induced immunity is repeated vaccination of entire populations. COVID-19 vaccination programmes targeting entire (adult) populations were estimated to be cost-saving at the time first and second vaccinations were introduced^9^, despite being financially and logistically resource intensive and often requiring diversion of other healthcare resources to deliver them. However, with an increasing proportion of the population having at least some level of immunity due to previous vaccinations and infections, combined with more recent SARS-CoV-2 variants being associated with lower risks of severe outcomes^10,11^, the potential reduced benefits and on-going high opportunity costs of vaccinating the entire population repeatedly should be carefully considered.

In contrast to vaccination, previous infection may offer longer-lasting protection^12^. Therefore, for low-risk populations, if the chance of harm from infection following initial vaccination is sufficiently small, frequently repeated vaccination paid from healthcare budgets may not be required and potentially even generate harm when considering the opportunity costs of not being able to spend this budget on other interventions that result in more gains in quality-adjusted life-years. For example, fourth (or fifth) vaccinations in the last quarter of 2022 are currently planned only for those aged 50y or older (or 75y or older or clinically vulnerable) in the UK, meaning natural infection will become the main immunological boosting mechanism for younger adults and children. However, natural infection could also bring risks such as exposure of vulnerable populations, complications including long COVID even in low-risk populations, and economic consequences to the society.

Whilst it is not yet possible to assess the impact of these fourth/fifth vaccinations, the substantial expansion of administration of third/booster mRNA vaccinations from 16 September 2021 in the United Kingdom (UK), in parallel with large numbers of breakthrough SARS-CoV-2 infections among those who had not yet received a third/booster vaccination, particularly with the emergence of the Omicron variants from mid November 2021, provides an opportunity to compare their impact on antibody responses.

We used data from the UK’s national COVID-19 Infection Survey (CIS), which is a large community-based representative survey randomly selecting private households across the UK, to investigate the duration of anti-trimeric spike IgG antibody responses following “breakthrough” infection vs third vaccination in those who had previously received two vaccinations but without evidence of prior infection. Antibody levels have been shown to be correlated with protection against infection in previous studies^7,13–15^, however, there is little information about antibody correlates of protection against the Omicron variants. We therefore also use CIS data to determine antibody-based correlates of protection against Omicron infections from 17 May 2022 onwards (predominantly BA.5 and BA.4 lineages) and use this to estimate how long infection is likely to be prevented after a third/booster vaccination or a breakthrough infection.

## Results

### Correlates of protection against Omicron BA.4/5 infection

We used data from 17 May 2022 to 12 September 2022 to determine the relationship between anti-spike antibody levels and protection from infection while Omicron BA.4/5 variants were dominant in the UK population. During this time period, of 19,311 sequenced infections, 13,097 (67.8%) were BA.5/sub-lineages, 2,924 (15.1%) BA.4/sub-lineages, 3,268 (16.9%) BA.2/sub-lineages, and 22 (0.1%) were other Omicron recombinants or Delta.

To determine correlates of protection against Omicron BA.4/5 infection and the effect of previous infection, the vaccinated population was divided into two groups, those without evidence of previous infection (62,146 participants, 106,653 visits) and vaccinated participants with evidence of previous infection (58,373 participants, 98,036 visits). Unvaccinated participants were excluded due to insufficient data (1,151 participants, 1,840 visits). Participants with previous infection were further divided into those with pre-Alpha/Alpha infection, with Delta/Omicron BA.1 infection (combined as effects similar, **Supplementary Figure 1**), and with Omicron BA.2 infection (**Supplementary Table 1**). Using logistic generalised additive models (GAMs) with PCR swab test results from each study visit as the outcome and the most recent antibody measurement obtained 21–59 days earlier, we found that protection against Omicron BA.4/5 infection increased with higher antibody levels in all groups; the increase was rapid for anti-spike IgG <2,000 BAU/mL and flattened after that. Higher antibody levels were needed to achieve the same level of protection in those without previous infection compared to those with previous infection. At the same antibody level, previous Delta/Omicron BA.1 infection afforded higher protection against Omicron BA.4/5 infection than a previous Pre-Alpha/Alpha infection. For example, antibody levels associated with 67% protection against infection for vaccinated participants aged 60y without previous infection, with Pre-Alpha/Alpha infection, and Delta/Omicron BA.1 infection were 1520 (95% confidence interval [CI] 1300-1840) BAU/mL, 1080 (95% CI 880-1320) BAU/mL, and 480 (95% CI 320-720) BAU/mL, respectively. A previous Omicron BA.2 infection had the highest protection against Omicron BA.4/5 infection, with over 80% of participants protected regardless of antibody levels (**Figure 1a**). Protection against moderate to high viral load infections (cycle threshold (Ct) values <30) and symptomatic infection was similar (**Figure 1b**,**c**).

**Figure 1.**
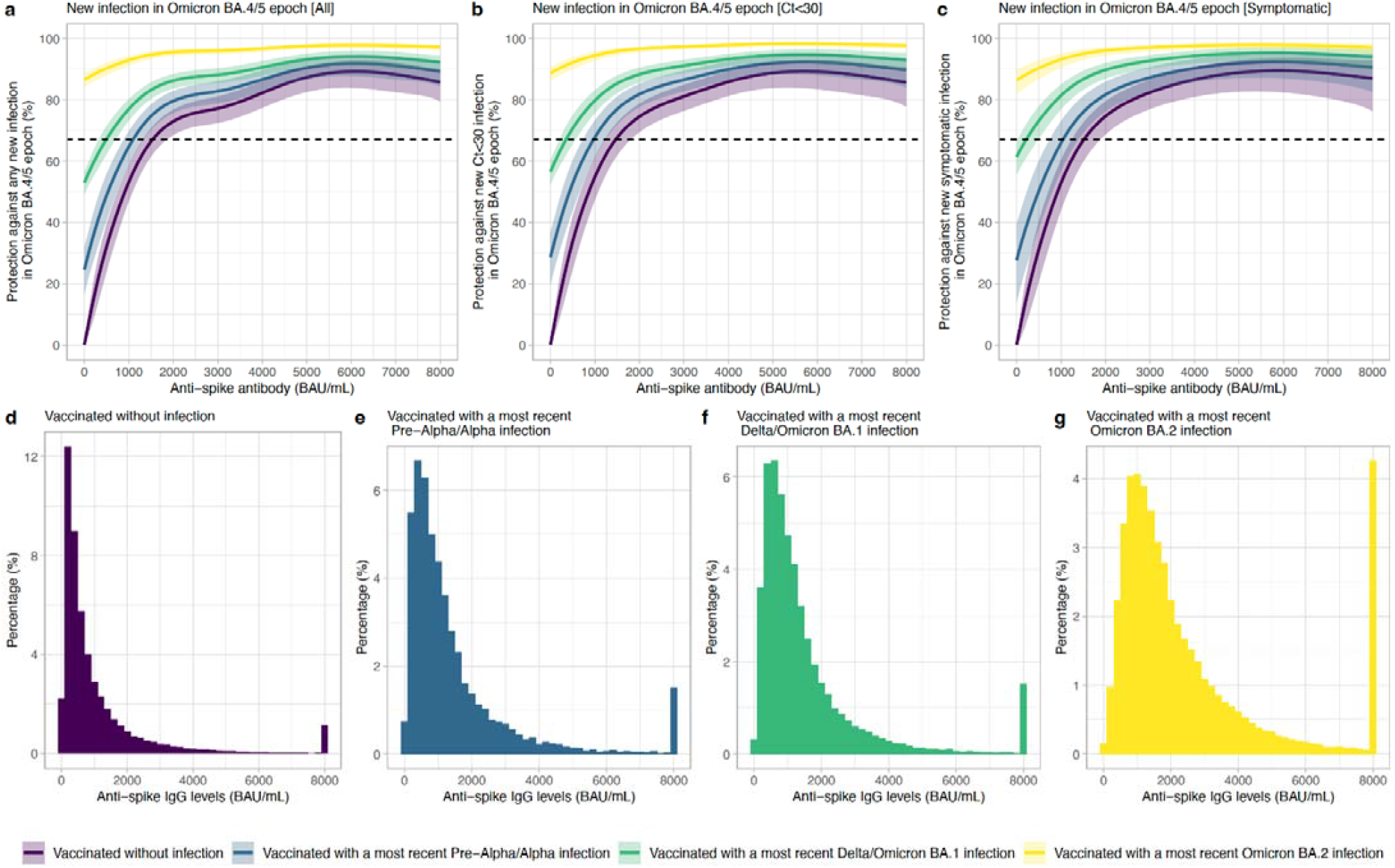
Association between anti-spike IgG levels and protection from SARS-CoV-2 infection using the most recent antibody measurement obtained 21–59 days before the current visit. **a**, Protection against any new infection in the Omicron BA.4/5 epoch. **b**, Protection against infection with a moderate to high viral load (Ct value < 30) in the Omicron BA.4/5 epoch. c, Protection against infection with self-reported symptoms in the Omicron BA.4/5 epoch. The 95% CIs are calculated by prediction ± 1.96 × standard error of the prediction. Four groups are investigated: vaccinated participants without evidence of prior infection, vaccinated participants with Pre-Alpha or Alpha infection, vaccinated participants with Delta or Omicron BA.1 infection, vaccinated participants with Omicron BA.2 infection. Distribution of the most recent anti-spike IgG measurements for the four population groups are shown in **d–g**.

Older participants, especially those ≥70y, required a higher antibody level to reach the same level of protection. For example, in those without previous infection the antibody levels associated with 67% protection were 1180-1520 BAU/mL for those aged 20-50y but were 2400 and 3200 BAU/mL at 75y and 80y. The differences were smaller between younger and older participants with a previous infection (**Supplementary Figure 2**). Time from the last vaccination/infection had a limited impact on the results, suggesting variant per se rather than time since infection was the main determinant of differences between those with previous Pre-Alpha/Alpha, Delta/Omicron BA.1 and BA.2 infections (**Supplementary Figure 3**).

### Antibody trajectories after third/booster vaccination and breakthrough infection

To estimate antibody trajectories, we used antibody measurements from 2 March 2021 to 12 September 2022. 154,149 participants aged ≥18y received two SARS-CoV-2 vaccinations with ChAdOx1 (with a 6-13 week dosing interval) or BNT162b2 (with a 3-13 week dosing interval) followed by a BNT162b2 or mRNA-1273 third/booster vaccination or a breakthrough infection (**Supplementary Table 2**). These included participants also had at least one antibody measurement after the second vaccination and no evidence of previous infection before the second vaccination.

We estimated rates of antibody decline from 21 days after the second vaccination and increases post-third/booster vaccination or infection using Bayesian piecewise linear interval-censored models. Different models were fitted for each primary course (ChAdOx1 or BNT162b2) and boosting event (BNT162b2 or mRNA-1273 third/booster vaccination or infection). We adjusted for age, sex, long-term health conditions, ethnicity (white vs non-white), working in healthcare, and time from second vaccination to booster/infection. Overall, the median age at third/booster vaccination or infection was 60y (Interquartile range [IQR] 50-69), 84,080 (54.5%) participants were female, and 147,712 (95.8%) reported white ethnicity. 43,175 (28.0%) reported having a long-term health condition, and 4,198 (2.7%) were healthcare workers. The median duration from the second vaccination to third/booster vaccination was 6 months and to infection was 5 months. The characteristics of those included in the antibody decline model from 42 days after the third/booster vaccination or infection were similar to the overall population (**Supplementary Table 3**).

As expected given associations between antibody levels and risk of infection^7^, antibody levels after the second vaccination were consistently lower in those who went on to become infected than those who received a third/booster vaccination (before infection) (**Figure 2**; orange vs blue/red lines), as well as being lower in those receiving a primary ChAdOx1 compared to a BNT162b2 course (dashed vs solid lines). Mean antibody levels were significantly boosted regardless of primary course or type of boosting event (**Figure 2**), to levels higher than that achieved after the second vaccination. The relatively lower antibody levels in those who received ChAdOX1 as a primary course were boosted by a greater degree than those who received a BNT162b2 primary course, such that post-booster both groups achieved similar antibody levels. mRNA-1273 boosters generated higher peak antibody levels than BNT162b2 boosters, but their subsequent waning was faster. Breakthrough infection boosted antibody levels to a similar level to the BNT162b2 booster, slightly lower than the mRNA-1273 booster. Estimated antibody declines after a booster/infection were generally faster following a ChAdOx1 primary course, compared to a BNT162b2 primary course. Following a ChAdOx1 primary course, estimated antibody declines were similar in participants following infection or a booster vaccination, for example, the half-life was 83 (95% credible interval [CrI] 63-118) days after infection compared to 78 (95% Crl 72-86) days with BNT162b2 booster and 63 (95% Crl 61-66) days with an mRNA-1273 booster for those aged 55y. Following a BNT162b2 primary course, the point estimate for decline after infection was slower than that after the booster, for example, the half-life was 141 (91-285 days) compared to 98 (91-106) days with a BNT162b2 booster and 93 (82-107) days with a mRNA-1273 booster for those aged 55y, although the credible intervals were wider due to smaller sample sizes (**Figure 2, Figure 3a,b**, **Supplementary Table 4**).

Younger individuals generally generated higher antibody levels 42 days post-booster vaccination than older individuals, except those who had BNT162b2 primary course and mRNA-1273 booster (**Figure 4**). Antibody levels were higher in individuals with a longer duration between the second vaccination and a breakthrough infection or BNT162b2 booster vaccination, but not an mRNA-1273 booster (**Figure 4**). There were relatively modest effects of other participant characteristics (**Supplementary Figure 4**).

**Figure 2.**
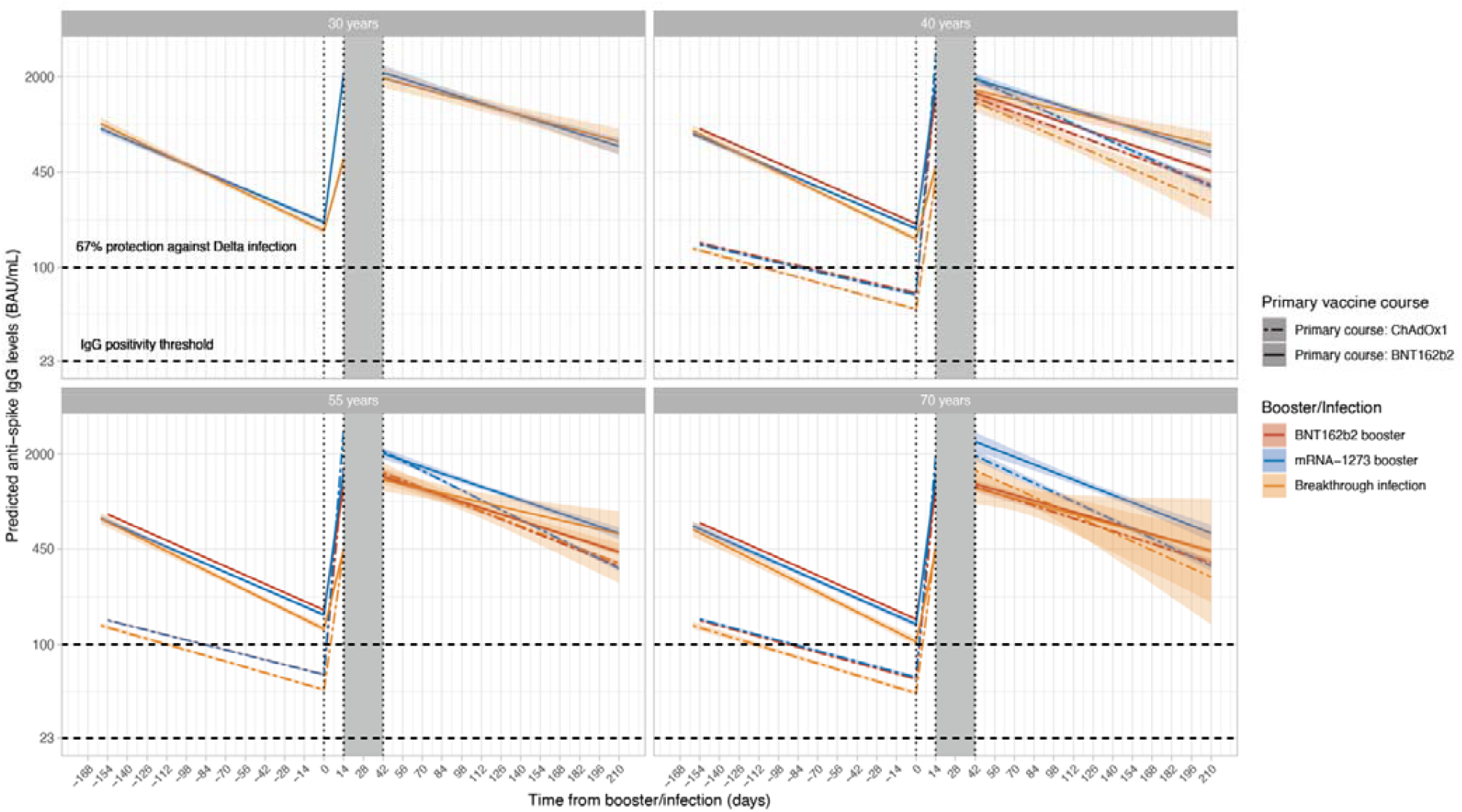
Posterior predicted mean trajectories (95%Crl) of anti-spike IgG levels from second vaccination through third/booster vaccination or infection using Bayesian linear mixed interval censored models. For each group, two separate models are fitted: 1) piecewise model on antibody decline after the second vaccination and subsequent increase after third/booster vaccination or infection; 2) antibody decline 42 days after the third/booster vaccination or infection. Shaded area between 14- and 42-days post third/booster vaccination or infection represents different timepoints individuals reach peak antibody levels. Models are adjusted for age, sex, ethnicity, time from second vaccination to booster/infection, long-term health conditions, and healthcare role. Plotted at the reference categories (female, white ethnicity, 6 months between second vaccination and booster/infection, not reporting a long-term health condition, not working in healthcare). Line types indicate the primary vaccine course. Line colours indicate the booster type or infection. Plots are separated by age (30y only estimated for those who had BNT162b2 as primary and were boosted by mRNA-1273 or infection due to low numbers in other groups). Predicted values are plotted on a log scale. Black dashed lines indicate the correlate for 67% protection against Delta variant (100 BAU/mL) and the threshold of IgG positivity (23 BAU/mL).

**Figure 3.**
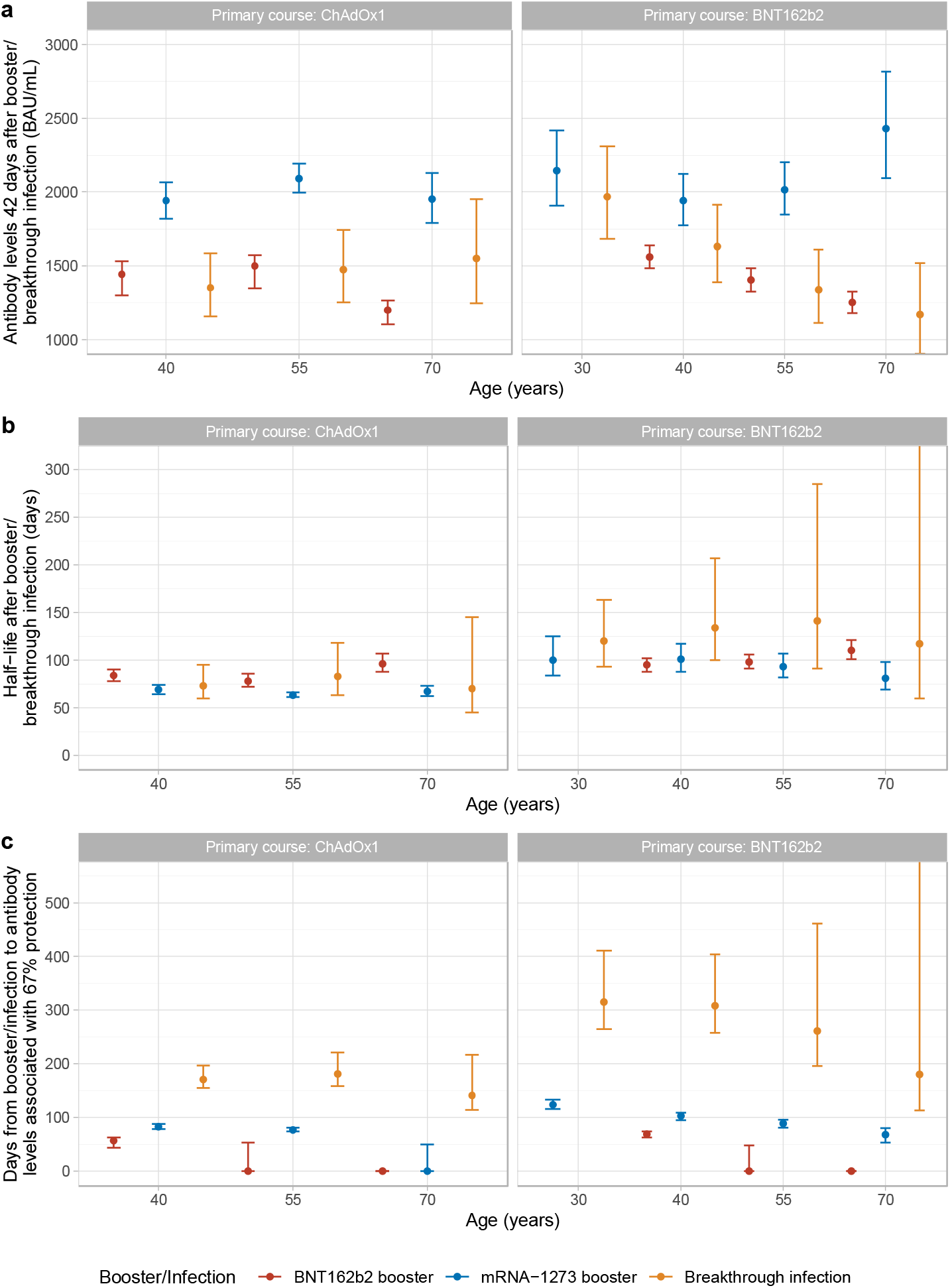
Comparisons of antibody levels 42 days post third/booster vaccination or infection, half-lives, and days from third/booster vaccination or infection to reaching antibody levels associated with 67% protection by primary vaccine course, third/booster vaccination or infection, and age. **a**, comparisons of antibody levels 42 days post third/booster vaccination or infection. **b**. comparisons of half-lives after third/booster vaccination or infection. **c**, comparisons of days from third/booster vaccination or infection to reaching antibody levels associated with 67% protection. Median values with 95% credible intervals are plotted. Numbers are shown in **Supplementary Table 3**. Plotted at the reference categories (female, white ethnicity, 6 months between second vaccination and booster/infection, not reporting a long-term health condition, not working in healthcare).

**Figure 4.**
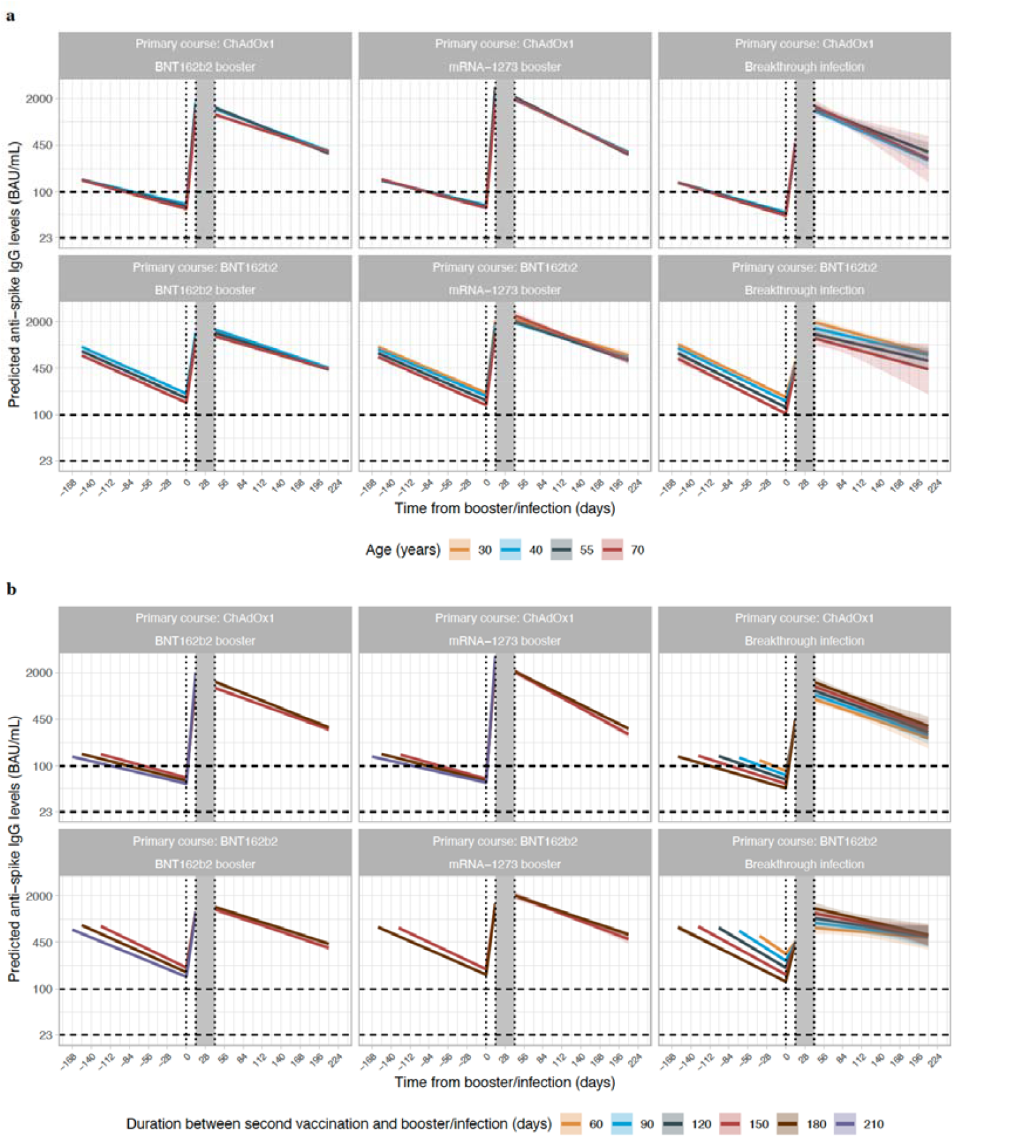
Posterior predicted mean trajectories (95%Crl) of anti-spike IgG levels from third/booster vaccination or infection. a, by age. b, by time from second vaccination to third/booster vaccination or infection. For each group, two separate models are fitted: 1) piecewise model on antibody decline after the second vaccination and subsequent increase after third/booster vaccination or infection; 2) antibody decline 42 days after the third/booster vaccination or infection. Shaded area between 14- and 42-days post third/booster vaccination or infection represents different timepoints individuals reach peak antibody levels. Models are adjusted for age, sex, ethnicity, time from second vaccination to booster/infection, long-term health conditions, and healthcare role. Plotted at the reference categories (female, white ethnicity, time from second vaccination to booster/infection 6 months, not reporting a long-term health condition, not working in healthcare). Plots are separated by primary vaccine courses and booster types or infection. Predicted values are plotted on a log scale. Black dashed lines indicate the correlate for 67% protection against Delta variant (100 BAU/mL) and the threshold of IgG positivity (23 BAU/mL).

There was no evidence of differences in antibody peak levels or half-lives across breakthrough infections before a booster with different variants (Delta or Omicron BA.1, the dominant variants causing breakthrough infections in the study) (**Supplementary Figure 5**).

### Protection from the third/booster vaccination and breakthrough infection against Omicron BA.4/5 infection

We combined our estimates of protection against infection by antibody level and of antibody declines to estimate the duration of protection against Omicron BA.4/5 infection.

We estimated the time from a third/booster vaccination or infection to mean antibody levels reaching levels associated with 67% protection against infection. The BNT162b2 booster did not provide this level of protection for participants aged >55y. For those aged 40y, antibody levels reached the threshold 60-70 days after the BNT162b2 booster, compared with around 80-100 days for participants receiving the mRNA-1273 booster. For those aged 70y, mRNA-1273 booster did not provide 67% protection at any point if they had received a ChAdOx1 primary course, compared with 60 days if they had received a BNT162b2 primary course. For participants with breakthrough infection, antibody levels associated with 67% protection lasted for 140-170 days with a ChAdOx1, and 180-315 days with a BNT162b2, primary course (**Figure 3c, Supplementary Table 4**). This is predominantly explained by the greater protection found at a given antibody level following infection compared to a booster, with some contribution from slower antibody waning following a BNT162b2 primary course.

We also estimated the proportion of participants with 67% protection at 42, 90, 180, 270, and 360 days from a third/booster vaccination or infection based on individual-level predictions, and assuming no further vaccination/infection. Following a ChAdOx1 primary course, 42 days after a BNT162b2 and mRNA-1273 booster, 60% and 100% of those aged 40-55y, and 20% and 90% of those aged 55-70y had antibody levels associated with ≥67% protection. However, no participant remained above this threshold level at 90 days. Following a BNT162b2 primary course, over 80% of those aged <55y with a BNT162b2 booster and nearly everyone with a mRNA-1273 booster were above the threshold level 42 days after the booster. At 90 days, almost every participant with a BNT162b2 booster fell below the level of 67% protection, while >90% of those aged <55y and 50% of those aged 55-70y with a mRNA-1273 booster remained above the threshold level. No participant remained above the threshold level at 180 days. For those with a breakthrough infection, nearly all participants aged <70y were above the threshold level at 42 and 90 days after infection. Nearly all participants who received a ChAdOx1 primary course had antibody levels below the 67% protection threshold level by 180 days. However, for those who received a BNT162b2 primary course, the percentage decreased over time but remained high (>70%) for those <55y at 270 days. The decrease was greater for older participants and almost every participant >55y did not maintain ≥67% protection by 270 days, although credible intervals were wide due to small numbers (**Figure 5**).

**Figure 5.**
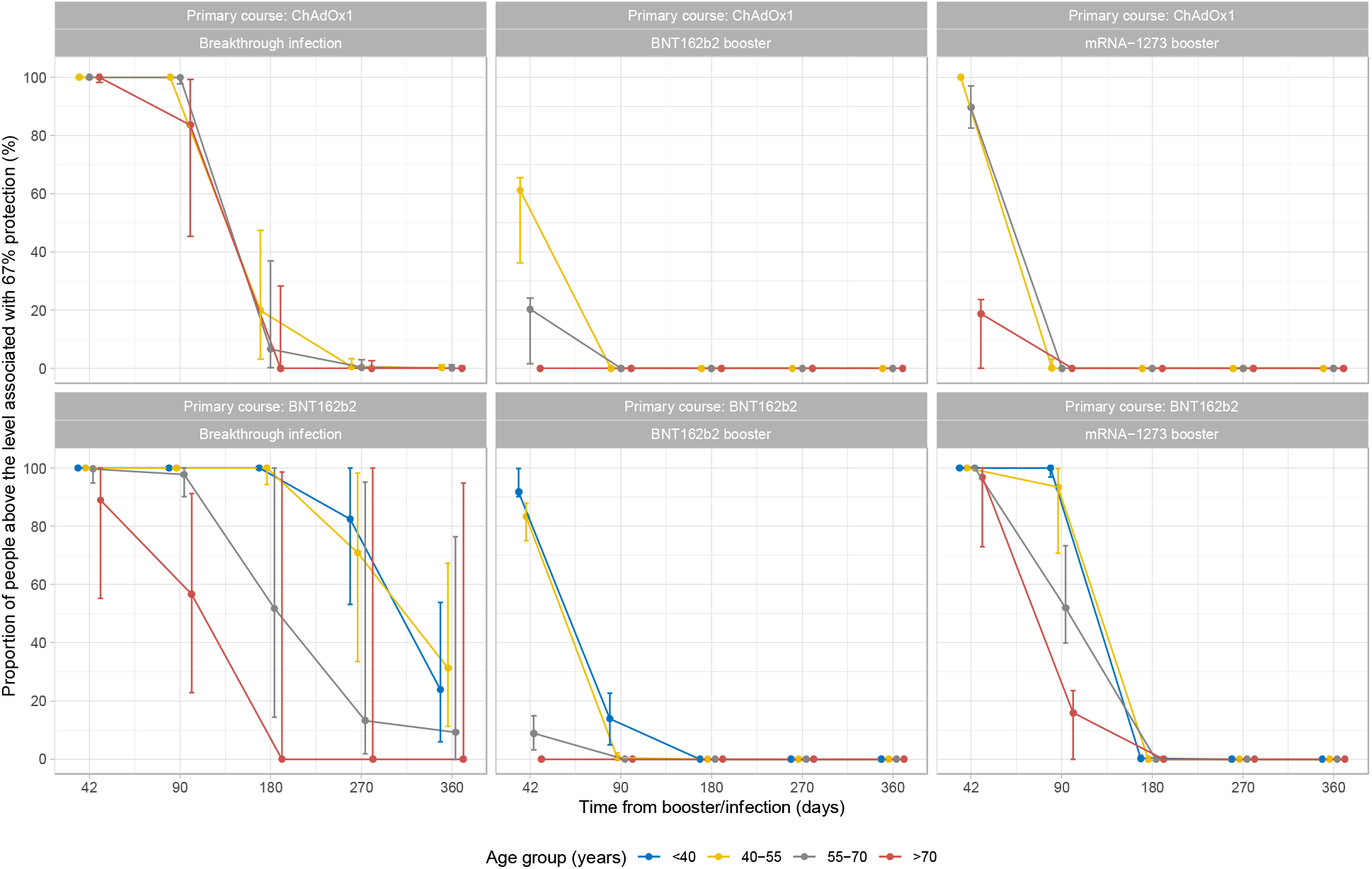
Proportion of participants above the anti-spike IgG antibody threshold level associated with 67% protection by time from third/booster vaccination or infection. Numbers of participants in each panel are [numbers in brackets represent <40, 40–55, 55-70 and >70 years]: ChAdOx1-Infection: n=4,123 [529, 2,138, 1,187, 269]; ChAdOx1-BNT162b2: n = 41,152 [1,295, 8,826, 19,232, 11,799]; ChAdOx1-mRNA-1273: n = 14,748 [738, 5,341, 7,859, 810]; BNT162b2-Infection: n = 1,834 [949, 436, 313, 136]; BNT162b2-BNT162b2: n = 24,749 [2,447, 3,613, 8,955, 9,734]; BNT162b2-mRNA-1273: n = 4,403 [1,791, 889, 1,409, 314]. ‘<40 year’ group is not plotted for ChAdOx1 primary course because the vast majority of those receiving ChAdOx1 were 40 years of age or older. Median values with 95% credible intervals are plotted.

To estimate a lower bound for levels of protection in the UK population, had further vaccination campaigns not taken place and the virus not been circulating to further boost antibody levels via breakthrough infections, we calculated the median protection levels over calendar time up to the end of 2022. In this scenario, the median level of protection in the UK population would be 50-60% among those who had a breakthrough infection and 5-15% among those who had triple vaccination without infection, with younger individuals having higher levels of protection than older individuals (**Figure 6**).

**Figure 6.**
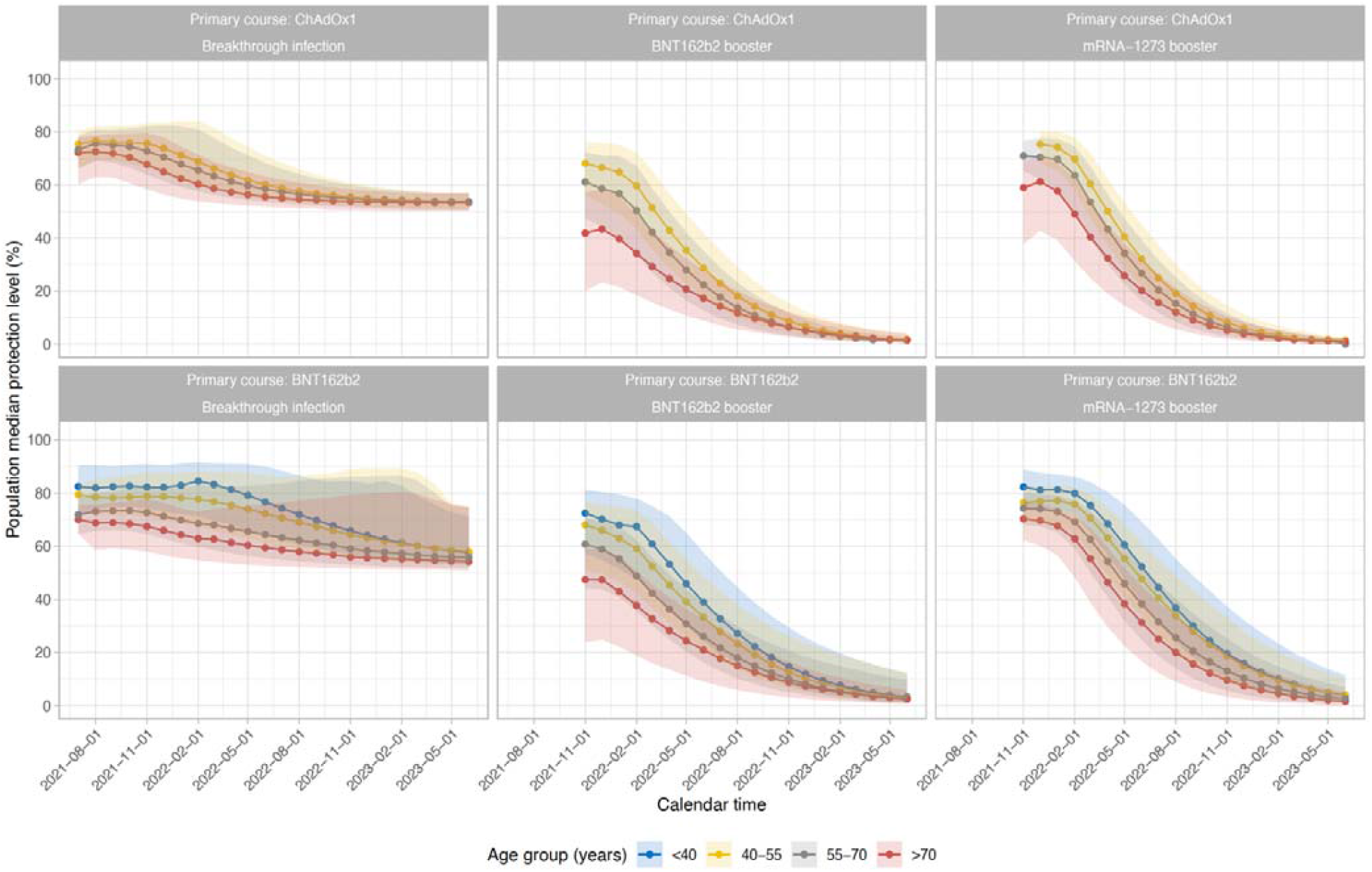
Median protection level by calendar time. Estimations were based on assumptions that participants did not receive another vaccination and were not infected after their third/booster vaccination or breakthrough infection. Numbers of participants in each panel are [numbers in brackets represent <40, 40–55, 55-70 and >70 years]: ChAdOx1-Infection: n=4,123 [529, 2,138, 1,187, 269]; ChAdOx1-BNT162b2: n = 41,152 [1,295, 8,826, 19,232, 11,799]; ChAdOx1-mRNA-1273: n = 14,748 [738, 5,341, 7,859, 810]; BNT162b2-Infection: n = 1,834 [949, 436, 313, 136]; BNT162b2-BNT162b2: n = 24,749 [2,447, 3,613, 8,955, 9,734]; BNT162b2-mRNA-1273: n = 4,403 [1,791, 889, 1,409, 314]. ‘<40 year’ group is not plotted for ChAdOx1 primary course because the vast majority of those receiving ChAdOx1 were 40 years of age or older. 95% credible interval is calculated from posterior simulations from the GAM model estimating correlates of protection and posterior predictions from the Bayesian linear mixed models estimating antibody levels.

## Discussion

In this UK population study, we found that breakthrough SARS-CoV-2 infection provided longer lasting protection against further infections than booster vaccinations, although both breakthrough infection and booster vaccination result in substantial increases in anti-spike IgG antibody levels, regardless of the third/booster vaccine type and the primary vaccine course. Breakthrough infections generated similar antibody levels to third/booster vaccinations, and the subsequent declines in antibody levels were similar or slightly slower than those after third/booster vaccinations. However, as antibody levels associated with the same level of protection level against new Omicron BA.4/5 infections were lower in those with a previous infection than those without, the duration of protection after a breakthrough infection was longer than after a third/booster vaccination.

Studies of neutralising antibodies have shown significantly reduced protection against Omicron compared to the wild type or preceding variants following two vaccinations^22–25^. We found that a third/booster vaccination substantially increased anti-spike IgG levels and led to higher antibody levels post-booster than post-second vaccination, similar to previous studies^16–18^. This at least partially explains the effectiveness of third/booster doses against Omicron infection compared to two doses^4,19^. Although antibody levels post-second vaccination were much lower after ChAdOx1 than BNT162b2, both groups in our study were boosted to a similar level, consistent with a previous VirusWatch study^20^, i.e. mRNA boosters significantly increased the relatively lower antibody levels induced by an adenovirus-vectored primary vaccination course.

We found that mRNA-1273 boosters resulted in higher antibody levels 42 days after the third/booster vaccination than BNT162b2. This is consistent with widely reported higher antibody levels from mRNA-1273 vs. BNT162b2 after second vaccinations^21–23^ and after a third/vaccination^24,25^. It is likely explained by mRNA-1273 delivering 100 μg mRNA per vaccine dose (50 μg for a booster dose), which is larger than BNT162b2 (30 μg mRNA). These results also potentially explain the higher vaccine effectiveness against new Omicron BA.1 infections reported 2-4 weeks after the mRNA-1273 booster (70.1-73.9%) than following the BNT162b2 booster (62.4-67.2%)^8^.

We found that breakthrough infections increased antibodies to similar levels to mRNA boosters, although the rate of increase was slower. Antibody declines after infection were similar to or slightly slower than after a booster, especially in those who had BNT162b2 primary course, suggesting a more sustained immune response post-infection than post-vaccination, consistent with our previous data^7,26^. Smaller studies have also found that neutralising activity was boosted by breakthrough infection after a second vaccination^22,27^ and was similar to that after three vaccinations^28^. Taken together with our data on correlates of protection, our results indicate that a breakthrough infection leads to longer-lasting immunity and thus offers more durable protection against future infections, both from the same and different variants. Sera from vaccinated individuals with breakthrough infections with pre-Omicron variants have been reported to cross-neutralise the Omicron variant, although less effectively than the delta variant^29^, and to a greater extent than sera from those without breakthrough infections^30–32^.

Multiple studies have shown antibody levels are a correlate of protection against infection, including several large studies or trials involving Alpha and Delta variant infections^7,13,15^, and Omicron BA.1 infections in a small healthcare worker cohort^33^. To our knowledge, our study is the first to show higher anti-spike IgG levels are associated with increased protection against Omicron BA.4/5 infection, the current predominant variants worldwide. We previously reported that the antibody level associated with 67% protection against new Delta infection was 100 BAU/mL in vaccinated individuals without prior infection^7^. Using the same 67% threshold, the antibody level required to provide the same level of protection against Omicron BA.4/5 infection was >1,000 BAU/mL in the same group, showing that much higher levels of antibodies against a wild-type trimeric spike antigen are needed to protect against new Omicron infection than new Delta infection, consistent with previous studies reporting lower neutralization of Omicron than Delta^34,35^. The level of protection associated with a given antibody level was strongly affected by infection and vaccination history, with vaccination without prior infection resulting in the lowest protection at a given antibody level compared to those with both vaccination and prior/breakthrough infection. Among those who had prior/breakthrough infection, the protection was highest for those who had a more recent variant of infection than those who had a preceding variant, but notably, there was no evidence of an effect of time since last infection on protection at a given antibody level having accounted for the most recent infecting variant. Populations with an Omicron BA.2 breakthrough infection were estimated to have >80% protection against an Omicron BA.4/5 reinfection.

Using 67% protection against infection as a threshold, protection was short-lived following three vaccinations in individuals without previous infection. The population-average time from third/booster vaccination to the level associated with 67% protection was no more than 70 days for the BNT162b2 booster and 125 days for the mRNA-1273 booster. The estimated duration was longer following two vaccinations and a breakthrough infection with Delta or Omicron BA.1, being 140-170 days with a ChAdOx1 primary course, and 180-315 days with a BNT162b2 primary course. This was partly because of slower antibody declines with a BNT162b2 primary course, but predominantly due to the lower antibody levels associated with 67% protection after breakthrough infections. The proportion of individuals above the 67% threshold showed similar patterns. At 180 days, no participant with a third/booster vaccination remained above the threshold, while all participants <55y with a BNT162b2 primary course and a breakthrough infection maintained 67% protection. This is consistent with previous studies in Qatar and Netherlands where higher protection was observed against the Omicron BA.1/BA.2 variant in people with both vaccination and previous infection than those who had only been vaccinated^36,37^.

Based on this cohort, which is broadly representative of the UK population, assuming participants did not have a further immune boosting event (vaccination or infection) after their third vaccination or breakthrough infection, the proportion of people who would remain protected at the 67% threshold in the UK would be <15% among those without a previous infection, and 50-60% among those with a previous infection by the end of 2022. To increase population immunity, a further booster vaccination would be helpful for those who have not had a SARS-CoV-2 infection (estimated from our cohort to be between 25-35% of those aged under 55y, **Supplementary Figure 6**), and those ≥55y with or without a previous infection. Currently, in the last quarter of 2022, a fourth vaccination is offered to those aged ≥50y in the UK (fifth for those aged ≥75y or clinically vulnerable). For the younger population, relatively robust immunity is likely to have already been acquired from a previous SARS-CoV-2 infection, and arguably the clinical risks from a new SARS-CoV-2 infection are much smaller^10,11,38^.

Given these results, providing risks of hospitalisation/death and onward transmission to at-risk groups remain acceptably low, breakthrough infections may be an efficient mechanism to maintain immunity in healthy younger individuals without clinical vulnerability. However, there would still be some risks associated with this approach, such as ongoing circulation of the variant that could put elderly and vulnerable populations at risk, and SARS-CoV-2 complications could still occur even in low-risk younger population. Ongoing infection might also cause economic or societal consequences even with low morbidity and mortality. Nevertheless, continuing with the widespread use of booster vaccination is associated with substantial costs, both the direct costs and the opportunity costs from diversion of healthcare. Taken together with the lower effectiveness of current vaccines against Omicron infection than against earlier variants, continuing vaccinating the whole population may have limited benefits. New vaccines with a higher effectiveness against Omicron variants or more sustained protection could be beneficial, but the current Omicron-specific vaccines only offer similar protection to existing booster vaccines^39^. Therefore, breakthrough infection could still be a reasonable immune-boosting strategy for subgroups of the population that have low risks of adverse consequences from infection.

Limitations of the study include the fact that we only measured anti-spike IgG and assumed that the antibody correlates of protection were constant over time; other immune mechanisms may also provide protection against infections, including T cell and memory-based responses. Neutralising antibody responses were not assayed in this study. We only measured antibody levels in a single assay and models for antibody responses to booster vaccination/infection included measurements from three different dilutions to cover varying ranges of observed values over time. However, we used interval censored methods to account for different censoring thresholds, and models for correlates of protection only used the highest dilution, in place when Omicron BA.4/5 infections dominated. We could not model participants with a primary vaccination course of mRNA-1273 due to insufficient data.

In summary, both third/booster vaccination and infection post-second vaccination significantly increased anti-spike IgG levels, regardless of the primary vaccine course. Breakthrough infections had at least as strong boosting effects, and antibody declines were similar or slightly slower after breakthrough infections than third/booster vaccinations. Based on the correlates of protection against new Omicron BA.4/5 infections, protection was lower and shorter after a third/booster vaccination, but higher and longer after breakthrough infection, especially among younger individuals. These results could inform future vaccine strategies against current and potentially future Omicron variants. Providing risks of hospitalisation/death, long-term complications, and onward transmission to at-risk groups remain acceptably low, breakthrough infections may be an efficient strategy to maintain immunity in healthy younger individuals without clinical vulnerability.

## Methods

### Participants and settings

The COVID-19 Infection Survey (CIS) (ISRCTN21086382, https://www.ndm.ox.ac.uk/covid-19/covid-19-infection-survey/protocol-and-information-sheets) is a large community-based survey with longitudinal follow-up, designed to be representative of the UK’s general population. Private households were randomly selected from address lists and previous surveys on a continuous basis for enrolment from 26 April 2020 through 31 January 2022 (when new recruitment was paused, although follow-up continued). After obtaining verbal agreement to participate, written informed consent was taken for individuals aged 2y and over by a study worker visiting each household. For those aged 2-15y, consent was provided by their parents or carers; those 10–15y also provided written assent. At the first visit, participants were asked for consent for optional follow-up visits every week for the next month and then monthly subsequently. The study received ethical approval from the South Central Berkshire B Research Ethics Committee (20/SC/0195).

At each visit, participants were asked about demographics, behaviours, work, and vaccination status. Combined nose and throat swabs were taken from all consenting household members for SARS-CoV-2 PCR testing. Blood samples were taken monthly for antibody testing from participants aged 16y and over in a randomly selected 10-20% of households. Household members of participants who tested positive on a nose and throat swab were also invited to provide blood monthly for follow-up visits. Details on the sampling design are provided elsewhere^40^. From April 2021, additional participants were invited to provide blood samples monthly to assess vaccine responses, based on a combination of random selection and prioritization of those in the study for the longest period (independent of swab test results).

### Vaccination data

Participants were asked about vaccination status at visits in the survey, including vaccination type, number of vaccinations, and vaccination dates. For participants from England, their vaccination data were also obtained from linkage to the National Immunisation Management Service (NIMS), which contains all individuals’ vaccination data in the English National Health Service COVID-19 vaccination program. We used records from the NIMS where available, otherwise used the self-reported data from the survey. There was good agreement between self-reported and administrative vaccination data (98% on type and 95% on date^41^).

### Laboratory testing

Combined nose and throat swabs were tested by PCR assays using the Thermo Fisher TaqPath SARS-CoV-2 assay at high-throughput national ‘Lighthouse’ laboratories in Glasgow and Milton Keynes (up until 8 February 2021). PCR outputs were analysed using UgenTec FastFinder 3.300.5, with an assay-specific algorithm and decision mechanism that allows conversion of amplification assay raw data into test results with minimal manual intervention. Samples are called positive if at least one single N-gene and/or ORF1ab are detected (although S-gene cycle threshold (Ct) values are determined, S-gene detection alone is not considered positive^40^) and PCR traces exhibiting an appropriate morphology. For contingency due to capacity issues, a small number of swabs were tested using endpoint PCR at the Rosalind Franklin laboratory (109,874 (12%) between 29 April and 12 September 2022, the period used in analyses of correlates of protection).

Venous or capillary blood samples were tested for SARS-CoV-2 antibody using an ELISA detecting anti-trimeric spike IgG developed by the University of Oxford^40,42^. Normalised results are reported in ng/ml of mAb45 monoclonal antibody equivalents. We used a commercialised CE-marked version of the assay, the Thermo Fisher OmniPATH 384 Combi SARS-CoV-2 IgG ELISA (Thermo Fisher Scientific), with the same antigen and colorimetric detection. mAb45 is the manufacturer-provided monoclonal antibody calibrant for this quantitative assay.

Antibodies were diluted at 1:50 for samples at the start of the survey. The dilution was changed to 1:400 from 28^th^ January 2022 and further changed to 1:1600 from 29^th^ April 2022 due to booster vaccinations and widespread Omicron infections causing saturated results. The 1:1600 dilution brought most test results within the dynamic range of the assay.

We calibrated the results of the Thermo Fisher OmniPATH assay into WHO international units (binding antibody unit, BAU/mL) using serial dilutions of the National Institute for Biological Standards and Control (NIBSC) Working Standard 21/234. A linear regression model fitted constrained to have an intercept of zero to convert mAB45 units in ng/ml to BAU/mL^7^:

BAU/mL = 0.559 * [mAb45 concentration in ng/mL]

The upper limit of quantification for the assay at 1:50, 1:400, 1:1600 dilutions were 450 BAU/mL, 1,360 BAU/mL, and 8,000 BAU/mL, respectively. Antibody measurements >450 BAU/mL under 1:50 dilution (251,471 observations, 22.7%), >1,360 BAU/mL under 1:400 dilution (46,346 observations, 4.2%), and >8,000 BAU/mL under 1:1600 dilution (1,696 observations, 0.2%) were truncated at 450 BAU/mL, 1,360 BAU/mL, and 8,000 BAU/mL, respectively. Antibody measurements <1 BAU/mL were truncated at 1 BAU/mL (194 measurements, 0.02%).

### Statistical analysis

#### Correlates of protection analysis

For the analysis of correlates of protection for new Omicron variants, we used data from study visits with antibody measured in the preceding 21-59 days from 17 May 2022 to 12 September 2022 inclusive due to the change of dilution on 29 April 2022 (therefore only including antibody measurements with 1:1600 dilution). The outcome was therefore mainly Omicron BA.4/5 infections (see Results). We included participants aged 18y and over, due to different recommendations regarding booster and primary vaccinations in younger participants. Analyses were based on visits. We grouped positive tests from the survey, the English national testing programme (national testing data were not available for Scotland, Wales, and Northern Ireland), and from self-reported positive swab tests into episodes^43^ because PCR-positive results might be observed at multiple visits after infection, and included only the first new PCR-positive visit in each infection episode. Visits occurring in the 21 days before and after each vaccination were excluded, as we have previously reported that infection rates change in the run-up to vaccination, reflecting structural bias (whereby those known to be positive defer planned vaccinations)^44^. We further included a positive anti-spike IgG result (≥23 BAU/mL) any time before the first vaccination or a self-reported positive test as a prior infection.

We used separate logistic generalised additive models (GAMs) for three outcomes: any positive PCR episode; a positive PCR episode with a moderate to high viral load (Ct value11<30); and a positive PCR episode with self-reported symptoms. As previously described^7^, we considered the effect of the most recent antibody measurement obtained 21–59 days before the current visit. The relationship between antibody levels and the outcome was modelled using thin-plate splines. Based on vaccination and infection history, we divided the population into vaccinated participants without evidence of previous infection (62,146 participants, 106,653 visits) and vaccinated participants with evidence of previous infection (58,373 participants, 98,036 visits). Unvaccinated participants were excluded due to insufficient data (1,151 participants, 1,840 visits). Vaccinated participants with a previous infection were further split by dominant variant at the earliest positive test in each infection episode as: Pre-Alpha (up to 6 December 2020, 3,490 participants, 5,945 visits), Alpha (07 December 2020-16 May 2021, 4,199 participants, 7,064 visits), Delta (17 May 2021-12 December 2021, 9,225 participants, 15,745 visits), Omicron BA.1 (13 December 2021-20 February 2022, 15,615 participants, 26,736 visits), and Omicron BA.2 (21 February 2022-05 June 2022, 22,079 participants, 38,223 visits) where dates were chosen as the first surveillance week (starting Monday) where >50% of positive tests matched the S-gene of the new variant (S-negative for Alpha, BA.1/4/5; S-positive for pre-Alpha, Delta and BA.2). Participants tested PCR-positive at 5,680 (5.3%) of visits without known previous infection and at 215 (3.6%), 258 (3.7%), 465 (3.0%), 529 (2.0%), and 201 (0.5%) visits after Pre-Alpha, Alpha, Delta, Omicron BA.1, or Omicron BA.2 infections respectively (**Supplementary Table 1**). We found that estimates from previous Pre-Alpha and Alpha infections, and previous Delta and Omicron BA.1 infections were similar, so we combined these visits to increase power (**Supplementary Table 1, Supplementary Figure 1**). The final model therefore consisted of four groups: vaccinated participants without evidence of infection before the current visit, vaccinated participants with a most recent Pre-Alpha or Alpha infection, vaccinated participants with a most recent Delta or Omicron BA.1 infection, vaccinated participants with a most recent Omicron BA.2 infection. Vaccinated participants with 1 (629 visits, 0.3%), 2 (7,657 visits, 3.7%), 3 (171,650 visits, 83.9%), or 4 (24,753 visits, 12.1%) doses of vaccines were grouped together, assuming that the effect of vaccination number was mediated through the resulting antibody levels. In the absence of an unvaccinated reference group, we compared these four groups to vaccinated participants without previous infection with a most recent anti-spike IgG measurement of 0 binding antibody units (BAU)/mL.

We adjusted for the following confounders in all models: vaccination and infection history (as above), time from last vaccination or infection, age in years, geographic area (9 regions in England, or the devolved administrations Wales, Scotland or Northern Ireland), rural/urban classification of home address, sex, ethnicity (white versus non-white), household size, multi-generational household, deprivation, presence of long-term health conditions, working in a care home, having a patient-facing role in health or social care, direct or indirect contact with a hospital or care home, and smoking status. Calendar time and age were included using a tensor spline, which was allowed to vary by region/country. We also included tensor spline interactions between antibody levels and age, and antibody levels and time since the last vaccination or infection, to examine the effects of age and time since the last event on the relationship between antibody and protection.

#### Antibody trajectory analysis

For the analysis of antibody trajectories, we included participants aged 18y and over who were eligible for third/booster vaccinations and had no evidence of previous infection. From 8^th^ December 2020 to 12^th^ September 2022, 259,561 participants received two vaccinations and had antibody measurements after the second vaccination. Of these, 32,810 participants were infected before the second vaccination and were excluded. 130,529 and 87,604 participants received two ChAdOx1 or BNT162b2 vaccinations for their primary course; 4,213 who received other vaccine types or mixed vaccine types were excluded. Among those who received two ChAdOx1 or BNT162b2 vaccinations, 194,679 received a third/booster vaccination (150,510 were boosted by BNT162b2, 43,257 were boosted by mRNA-1273). 912 received other vaccine types as a booster and were also excluded, and 25,544 had breakthrough infection post-second vaccination.

SARS-CoV-2 infection was defined as for the correlates analysis, namely as the earliest of a PCR-positive swab test in the survey, a positive swab test in the English national testing programme (national testing data were not available for Scotland, Wales, and Northern Ireland), a self-reported positive swab test or a positive anti-spike IgG result (≥23 BAU/mL) any time before the first vaccination. Antibody measurements after a third/booster vaccination that happened after a post-second vaccination infection, and antibody measurements after infection that happened post-booster were excluded from the analyses. We limited the analyses to those whose primary vaccination course was homologous ChAdOx1 (with a dosing interval 6-13 weeks) or BNT162b2 (with a dosing interval 3-13 weeks). We excluded a small number of participants who were non-responders after the second vaccination, defined previously^7^ as all antibody measurements being <16 BAU/mL after the second vaccination and having at least one antibody measurement 21 days after the second vaccination (N=416 excluded for ChAdOx1, N=174 excluded for BNT162b2). Age was truncated at 85y in all analyses to reduce the influence of outliers (1% of the population).

We used Bayesian linear mixed interval-censored models to estimate antibody levels and the effects of covariates on changes in antibody levels over time. Models were built separately by primary vaccine course (ChAdOx1 or BNT162b2) and booster type (BNT162b2 or mRNA-1273) or infection. Analyses of primary ChAdOx1 vaccinations included 64,940, 21,960, and 10,830 participants boosted by BNT162b2, mRNA-1273 and infection, respectively. Analyses of primary BNT162b2 vaccinations included 44,197, 7,248, and 4,974 participants boosted by BNT162b2, mRNA-1273, and infection, respectively. Antibody trajectory consisted of three slopes: 1) the decline (waning) from 21 days post-second vaccination to the third/booster vaccination or infection at t=0; 2) the increase from the third/booster vaccination or infection to the peak; 3) the decline (waning) after the peak post-booster or infection. Because there were variation in the time taken to reach peak antibody levels following different boosters/infection and in different study participants, the choice of the peak position could influence the estimate of the half-life post third/booster vaccination or infection, we separated the trajectories and built two models for each group: 1) a piecewise model: from 21 days post-second vaccination to 14 days post-third/booster vaccination or infection; 2) a decline model: from 42 days post third/booster vaccination or infection to better estimate the antibody decline. For the antibody declines, we excluded measurements taken after the 90^th^ percentile of the observed t>0 time points in each model since these outlying observations could have a strong and undue influence on the final decline after the peak post-booster/infection. The overall 90^th^ threshold still left some outliers for older age groups who had a breakthrough infection, so we further excluded measurements outside the specific 90^th^ percentile in these groups (**Supplementary Table 5**).

Population-level fixed effects, individual-level random effects for intercept and slopes, and correlation between random effects were included in all models. The outcome was modelled on the log2 scale and right-censored at 450, 1,360, and 8,000 BAU/mL for measurements in 1:50, 1:400, and 1:1600 dilution, respectively, reflecting truncation of IgG values at the upper limit of quantification (i.e. all measurements truncated to 450 BAU/mL in 1:50 dilution were considered to be >450 BAU/mL in analyses, same for 1:400 and 1:1600 dilutions). For the decline model, we only used antibody data at 1:400 and 1:1600 dilution to avoid the influence from a high proportion of antibody measurements censored at 1:50 dilution (>80%).

To examine non-linearity in antibody declines after third/booster vaccination or infection, especially the assumption that the rate of antibody decline would flatten, we additionally fitted a model using three-knot splines for time (knots placed at 10th, 50th and 90th of included time points) and compared with the linear models for each group. For all six groups, the estimated trajectories were similar, so we retained the log-linear model for analysis (**Supplementary Figure 7**).

Multivariable models included the effect of age, time from the second vaccination to the third/booster vaccination or infection, sex, ethnicity (white vs non-white due to small numbers in the latter), reporting having a long-term health condition, and reporting working in healthcare, each on the intercept (main effect) and on the slope (interaction). We used a 3-knot natural cubic spline for age (knots placed at 10^th^, 50^th^, and 90^th^ percentile of unique integer ages) to allow non-linear effects with antibody rising and waning post third/booster vaccination or infection. We restricted the range of time from the second vaccination to the third/booster vaccination or infection between the 10^th^ and 90^th^ percentiles. For infection models, we also examined the impact of infection type (Delta or Omicron BA.1) on antibody levels post-infection. For this analysis, infection type was defined primarily by sequencing data from CIS. If sequencing data was not available, Delta was defined as the first infection date in the most recent infection episode occurring from 10 April 2021 to 16 January 2022 and having at least one S-gene positive test (ORF1ab+N+S or ORF1ab+S or N+S) across the infection episode, Omicron BA.1 was defined as the first infection date in the most recent episode being after 29^th^ November 2021 and not having any S-gene positive tests. If gene positivity was not available (primarily infections from the national testing programme), infections that happened between 14^th^ June 2021 and 23^rd^ November 2021 were considered as Delta and those that happened after 20^th^ December 2021 were considered as Omicron BA.1. Infections with other variants or unknown variants were excluded from this model.

For each Bayesian linear mixed model, weakly informative priors were used (**Supplementary Table 6**). Six chains were run per model with 4,000 iterations and a warm-up period of 2,000 iterations to ensure convergence, which was confirmed visually and by ensuring the Gelman-Rubin statistic was <1.05 (**Supplementary Table 7**). 95% credible intervals were calculated using the highest posterior density intervals.

#### Estimation of protection

We combined our estimates of protection against infection by antibody level and of antibody declines to estimate the duration of protection against Omicron BA.4/5 infection. For those with a BNT162b2 or mRNA-1273 booster, we used estimates from the ‘vaccinated participants without previous infection’ group from the correlates model. For those with a breakthrough infection, since most earlier infections (>95%) were Delta and Omicron BA.1 (**Supplementary Table 3**), we used estimates from the ‘vaccinated participants with a most recent Delta or Omicron BA.1 infection’ group.

To estimate the actual population-level protection by calendar time, we combined results from the correlates of protection models and antibody trajectory models to get the population-level estimates. We used Metropolis Hastings posterior sampling to generate 100 sets of model coefficients from the correlates of protection models and extracted 100 draws from the posterior predictions from the antibody trajectory models, resulting in 10,000 predictions of actual protection level for each participant at each antibody level. Median protection level and 95% credible intervals were calculated by age group and calendar time. Protection estimations were based on assumptions that participants did not have previous infection before the first vaccination, did not receive another vaccination and were not infected after their third/booster vaccination or breakthrough infection.

Raw data were processed using Stata MP 16. All analyses were performed in R 3.6 using the following packages: tidyverse (version 1.3.0), mgcv (version 1.8-31), cenGAM (version 0.5.3), brms (version 2.14.0), splines (version 3.6.1), ggeffects (version 0.14.3), arsenal (version 3.4.0), cowplot (version 1.1.0), bayesplot (version 1.7.2).

## Data Availability

Data are still being collected for the COVID-19 Infection Survey. De-identified study data are available for access by accredited researchers in the ONS Secure Research Service (SRS) for accredited research purposes under part 5, chapter 5 of the Digital Economy Act 2017. Individuals can apply to be an accredited researcher using the short form on https://researchaccreditationservice.ons.gov.uk/ons/ONS_registration.ofml. Accreditation requires completion of a short free course on accessing the SRS. To request access to data in the SRS, researchers must submit a research project application for accreditation in the Research Accreditation Service (RAS). Research project applications are considered by the project team and the Research Accreditation Panel (RAP) established by the UK Statistics Authority at regular meetings. Project application example guidance and an exemplar of a research project application are available. A complete record of accredited researchers and their projects is published on the UK Statistics Authority website to ensure transparency of access to research data. For further information about accreditation, contact or visit the SRS website.

## Code availability

A copy of the analysis code is available at https://github.com/jiaweioxford/COVID19_booster_infection..

## Acknowledgements

We are grateful for the support of all COVID-19 Infection Survey participants.

This study is funded by the Department of Health and Social Care with in-kind support from the Welsh Government, the Department of Health on behalf of the Northern Ireland Government and the Scottish Government. JW is supported by University of Oxford and the China Scholarship Council. ASW, TEAP, NS, DE, KBP are supported by the National Institute for Health Research Health Protection Research Unit (NIHR HPRU) in Healthcare Associated Infections and Antimicrobial Resistance at the University of Oxford in partnership with the UK Health Security Agency (UK HSA) (NIHR200915). ASW and TEAP are also supported by the NIHR Oxford Biomedical Research Centre. KBP is also supported by the Huo Family Foundation. ASW is also supported by core support from the Medical Research Council UK to the MRC Clinical Trials Unit [MC_UU_12023/22] and is an NIHR Senior Investigator. PCM is funded by Wellcome (intermediate fellowship, grant ref 110110/Z/15/Z) and holds an NIHR Oxford BRC Senior Fellowship award. DWE is supported by a Robertson Fellowship and an NIHR Oxford BRC Senior Fellowship. NS is an Oxford Martin Fellow and holds an NIHR Oxford BRC Senior Fellowship. The views expressed are those of the authors and not necessarily those of the National Health Service, NIHR, Department of Health, or UKHSA.

## The COVID-19 Infection Survey team

Tina Thomas^9^, Daniel Ayoubkhani^9^, Russell Black^9^, Antonio Felton^9^, Megan Crees^9^, Joel Jones^9^, Lina Lloyd^9^, Esther Sutherland^9^, Emma Pritchard^1^, Karina-Doris Vihta^1^, George Doherty^1^, James Kavanagh^1^, Kevin K. Chau^1^, Stephanie B. Hatch^1^, Daniel Ebner^1^, Lucas Martins Ferreira^1^, Thomas Christott^1^, Wanwisa Dejnirattisai^1^, Juthathip Mongkolsapaya^1^, Sarah Cameron^1^, Phoebe Tamblin-Hopper^1^, Magda Wolna^1^, Rachael Brown^1^, Richard Cornall^1^, Gavin Screaton^1^, Katrina Lythgoe^2^, David Bonsall^2^, Tanya Golubchik^2^, Helen Fryer^2^, Stuart Cox^15^, Kevin Paddon^15^, Tim James^15^, Thomas House^16^, Julie Robotham^17^, Paul Birrell^17^, Helena Jordan^18^, Tim Sheppard^18^, Graham Athey^18^, Dan Moody^18^, Leigh Curry^18^, Pamela Brereton^18^, Ian Jarvis^19^, Anna Godsmark^19^, George Morris^19^, Bobby Mallick^19^, Phil Eeles^19^, Jodie Hay^20^, Harper VanSteenhouse^20^, Jessica Lee^21^, Sean White^22^, Tim Evans^22^, Lisa Bloemberg^22^, Katie Allison^23^, Anouska Pandya^23^, Sophie Davis^23^, David I Conway^24^, Margaret MacLeod^24^, Chris Cunningham^24^

^15^Oxford University Hospitals NHS Foundation Trust, Oxford, UK

^16^University of Manchester, Manchester, UK

^17^Health Improvement Directorate, Public Health England, London, UK

^18^IQVIA, London, UK

^19^National Biocentre, Milton Keynes, UK.

^20^Glasgow Lighthouse Laboratory, London, UK

^21^Department of Health and Social Care, London, UK

^22^Welsh Government, Cardiff, UK

^23^Scottish Government, Edinburgh, UK

^24^Public Health Scotland, Edinburgh, UK

## Author Contributions

The study was designed and planned by ASW, JF, JB, JN, ID and KBP, and is being conducted by ASW, RS, DC, ER, AH, BM, TEAP, PCM, NS, SH, EYJ, DIS, DWC and DWE. This specific analysis was designed by JW, DWE, ASW, and KBP. JW contributed to the statistical analysis of the survey data. JW, DWE, KBP and ASW drafted the manuscript and all authors contributed to interpretation of the data and results and revised the manuscript. DWE, KBP, and ASW contributed equally. All authors approved the final version of the manuscript.

