## supplementary for "Correlates of protection against SARS-CoV-2 Omicron variant and anti-spike antibody responses after a third/booster vaccination or breakthrough infection in the UK general population"

### Supplementary Tables and Figures

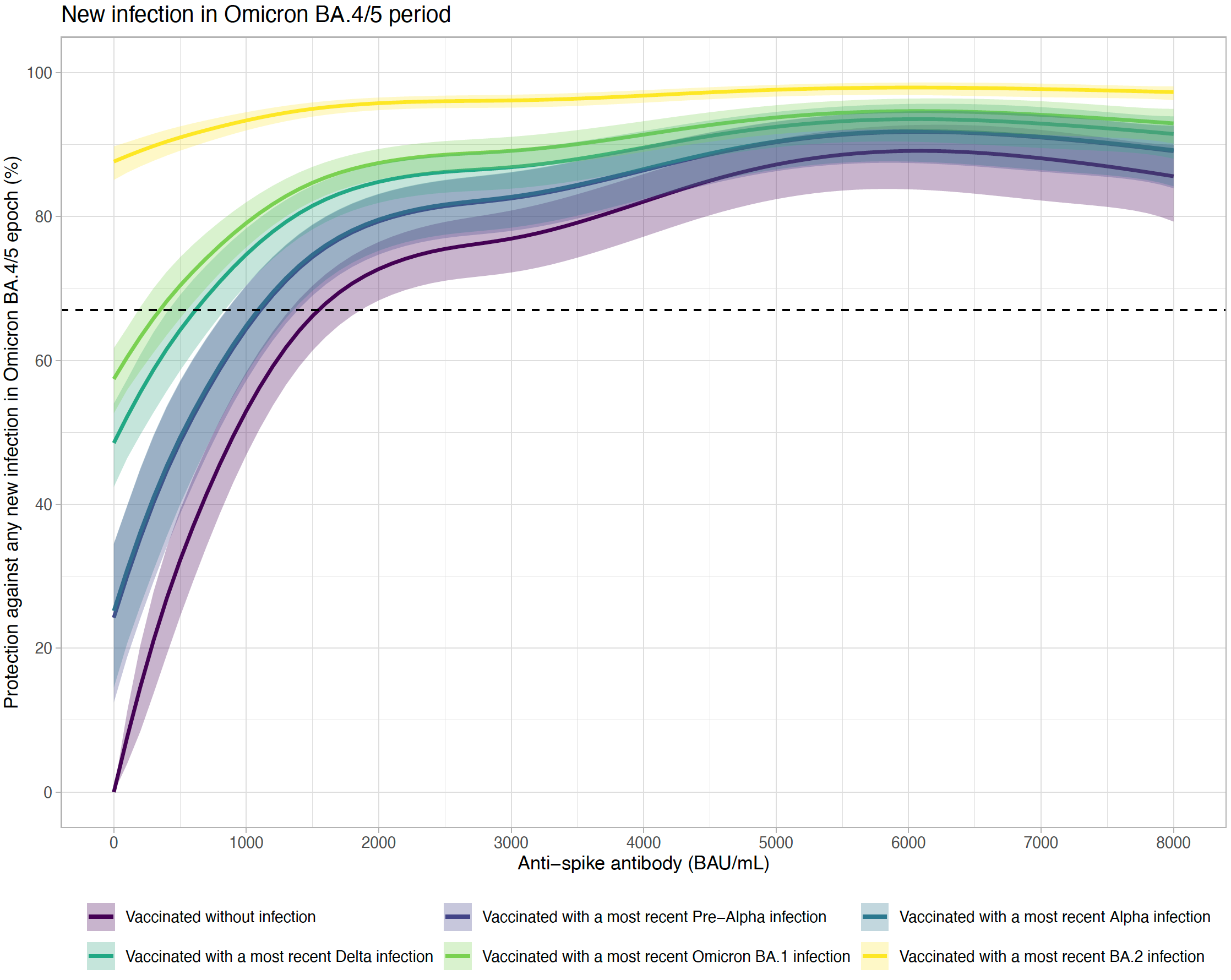

**Supplementary Figure 1. Association between anti-spike IgG levels and protection from SARS-CoV-2 infection using the most recent antibody measurement obtained 21–59 days before the current visit by infection variant.** The 95% CIs are calculated by prediction ± 1.96 × standard error of the prediction. From the plot, estimates on Pre-Alpha and Alpha infection, and Delta and Omicron BA.1 infection are similar. Therefore, Pre-Alpha and Alpha infection, and Delta and Omicron BA.1 infection are combined in the main model to increase power.

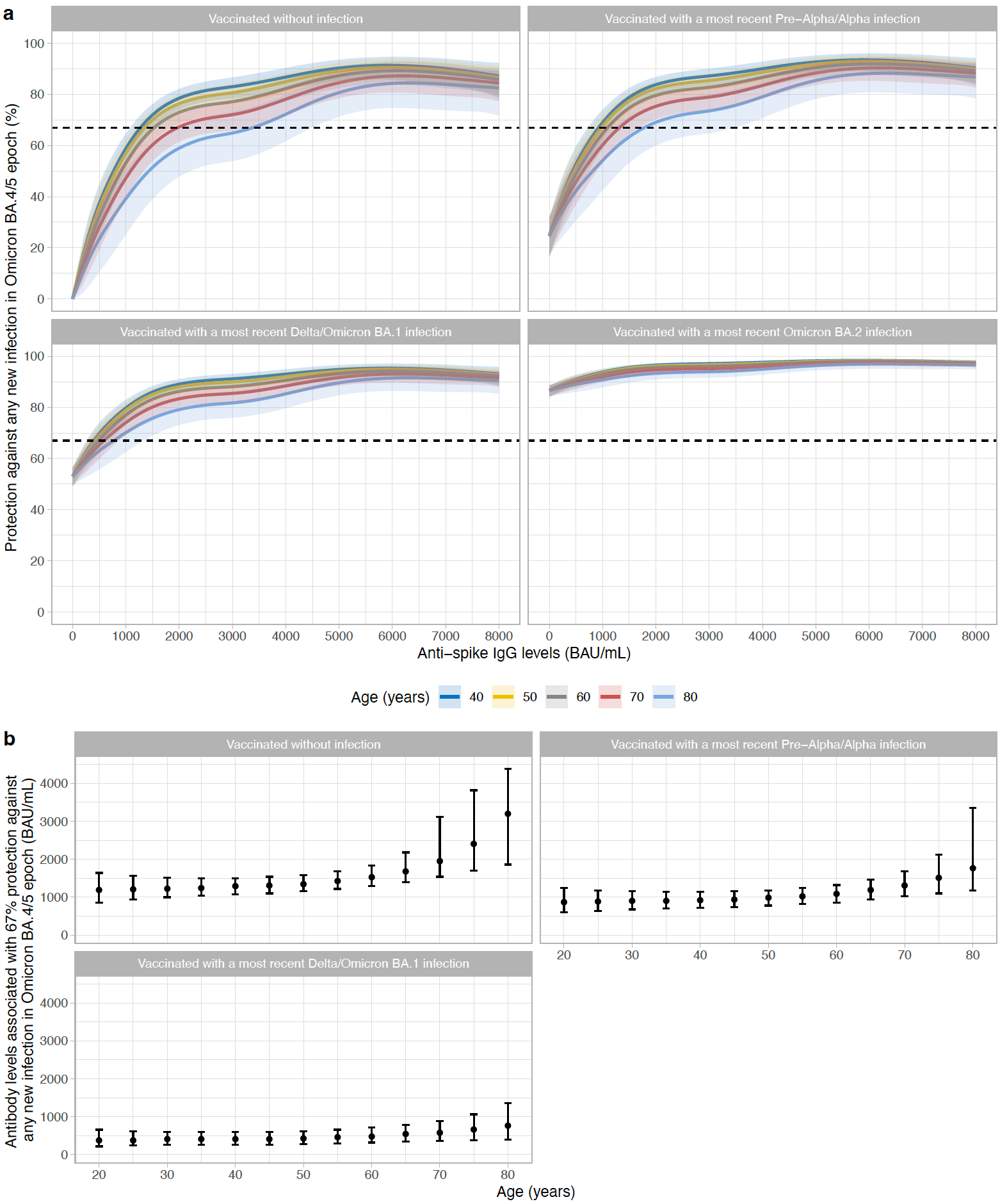

**Supplementary Figure 2. Age effects on correlates of protection. a,** Association between anti-spike IgG levels and protection from SARS-CoV-2 infection by age. Colour represents ages. Dotted line represents 67% protection. Four groups are investigated: vaccinated participants without evidence of prior infection, vaccinated participants with a most recent Pre-Alpha or Alpha infection, vaccinated participants with a most recent Delta or Omicron BA.1 infection, vaccinated participants with a most recent Omicron BA.2 infection. **b,** comparison of antibody levels associated with 67% protection against infection by age. The 95% CIs are calculated by prediction ± 1.96 × standard error of the prediction.

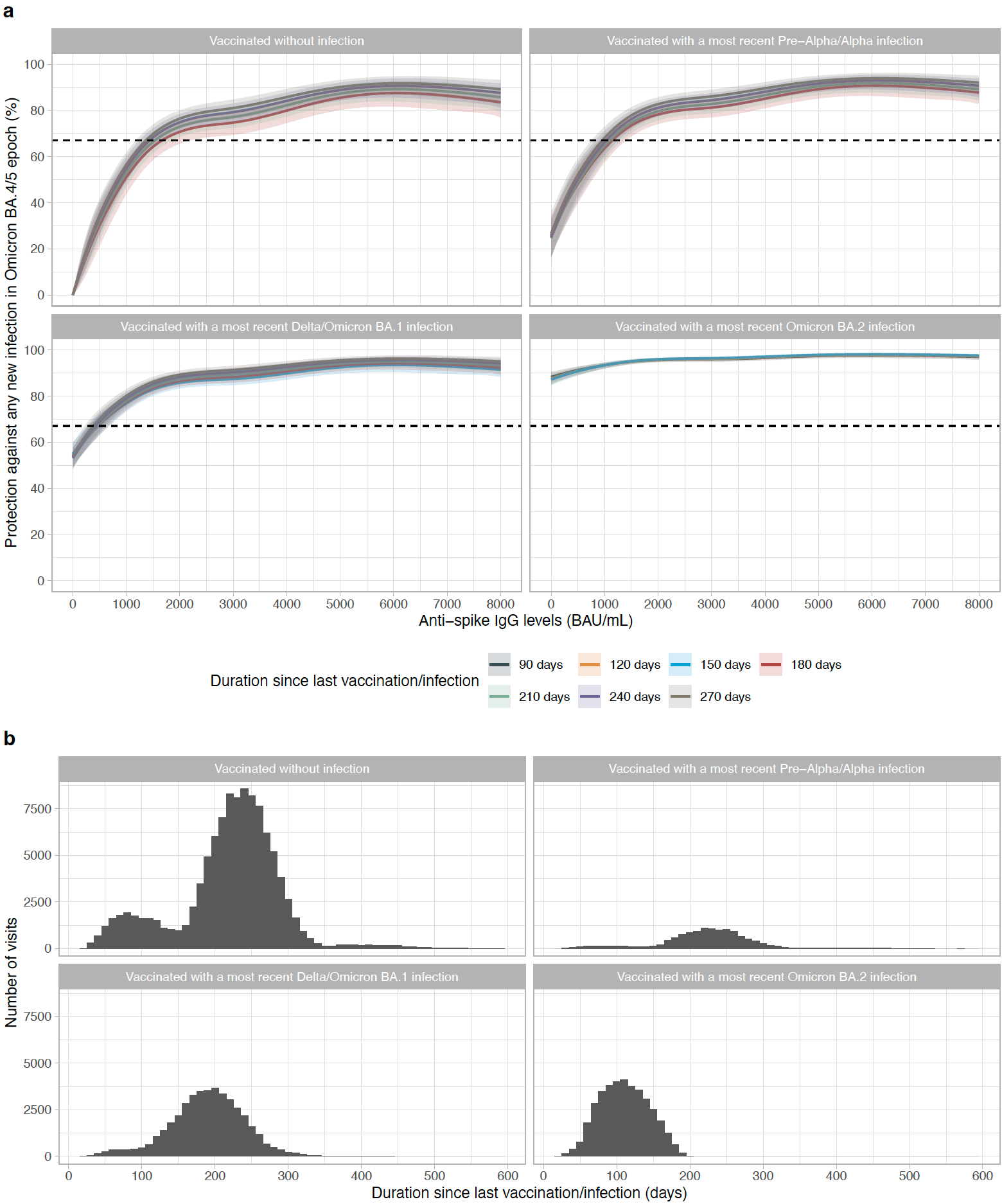

**Supplementary Figure 3. Effects from time since last vaccination or infection on correlates of protection. a,** Association between anti-spike IgG levels and protection from SARS-CoV-2 infection by duration since last vaccination or infection to the current visit. Colour represents duration since last vaccination or infection. Dotted line represents 67% protection. Four groups are investigated: vaccinated participants without evidence of prior infection, vaccinated participants with a most recent Pre-Alpha or Alpha infection, vaccinated participants with a most recent Delta or Omicron BA.1 infection, vaccinated participants with a most recent Omicron BA.2 infection. The 95% CIs are calculated by prediction ± 1.96 × standard error of the prediction. **b,** Distribution of duration from last vaccination or infection to the current visit by group. The median duration was 230, 230, 190, 110 days for the four groups.

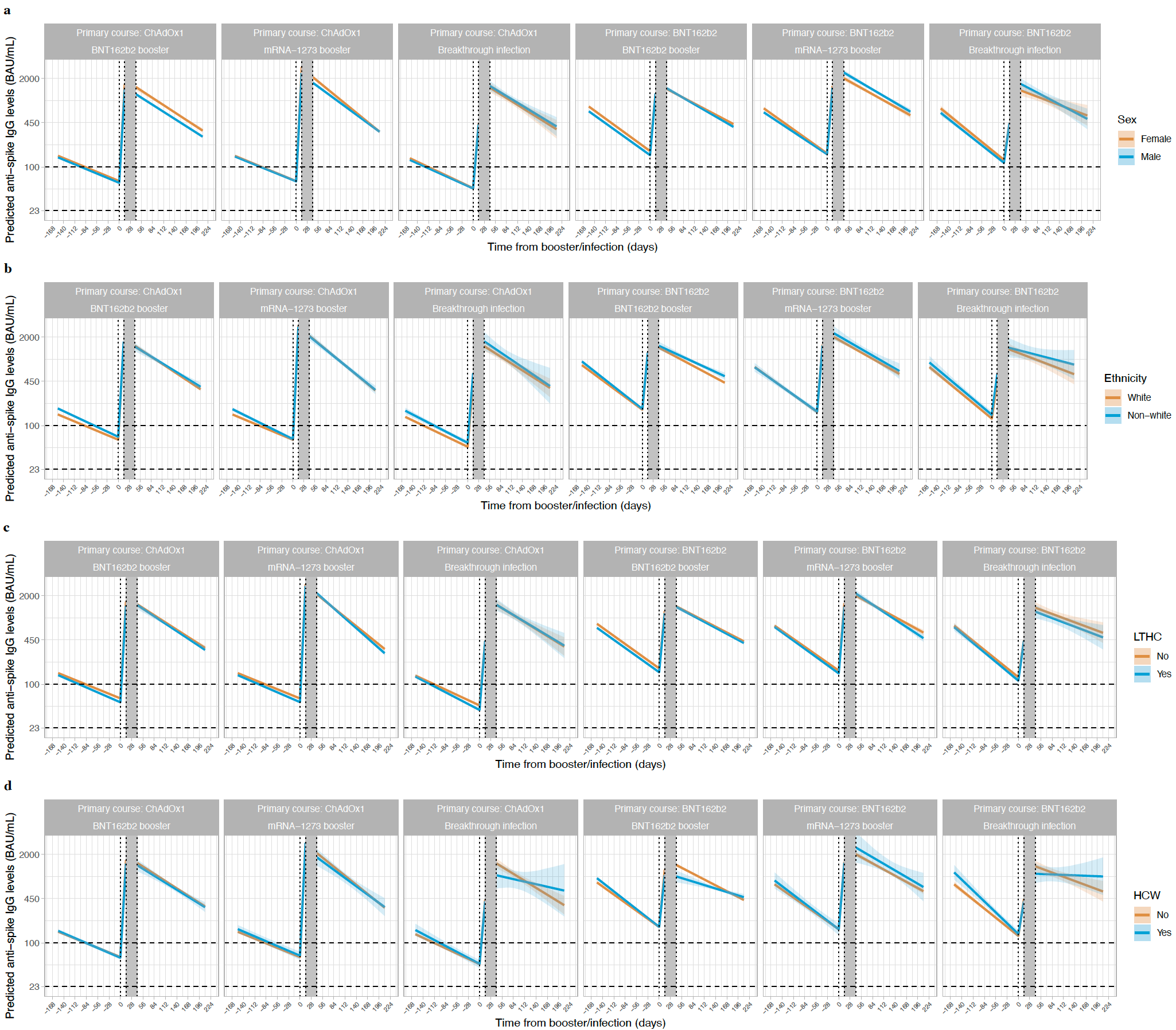

**Supplementary Figure 4. Posterior predicted mean trajectories (95%Crl) of anti-spike IgG levels from third/booster vaccination or infection by sex, ethnicity, long-term health condition, and healthcare role.** Models are adjusted for age, sex, ethnicity, time from second vaccination to booster/infection, long-term health conditions, and healthcare role. Plotted at the reference categories (55y, 6 months from second vaccination to booster/infection, female, white ethnicity, not reporting long-term health condition, not working in healthcare). Plots are separated by primary vaccine course and booster types or infection. Predicted values are plotted on a log scale. Black dashed lines indicate the correlate for 67% protection against Delta variant (100 BAU/mL) and the threshold of IgG positivity (23 BAU/mL).

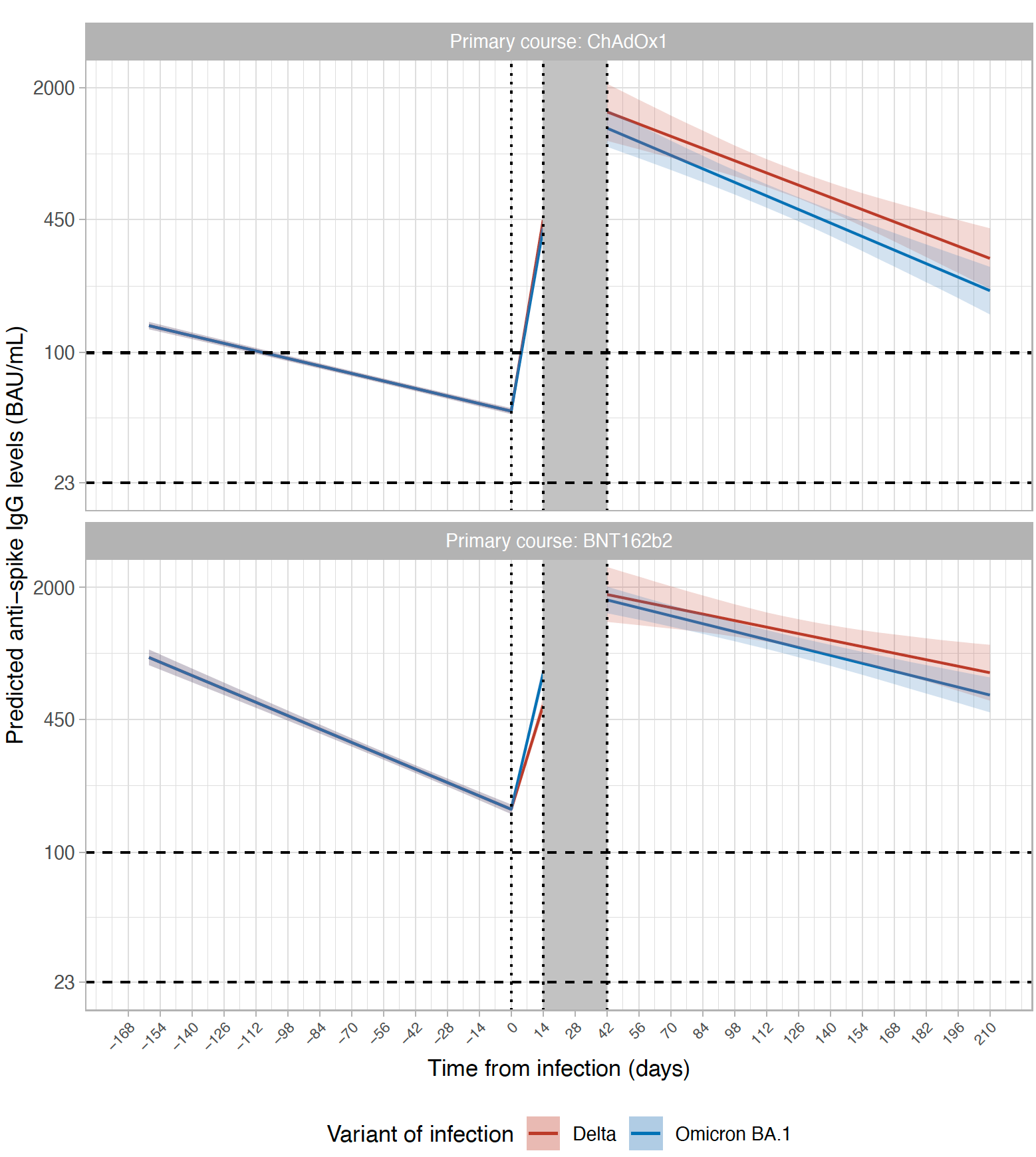

**Supplementary Figure 5. Effects of SARS-CoV-2 variant (Delta vs Omicron BA.1) on antibody responses after infection.** Plotted by primary vaccine course (ChAdOX1 or BNT162b2). Plotted at the reference categories (40y, 6 months from second vaccination to booster/infection, female, white ethnicity, not reporting long-term health condition, not working in healthcare). Black dashed lines indicate the correlate for 67% protection against Delta variant (100 BAU/mL) and the threshold of IgG positivity (23 BAU/mL). Shaded area between 14- and 42-days post infection represents different timepoints individuals reach peak antibody levels. No differences on antibody levels and subsequent decline by infection variant.

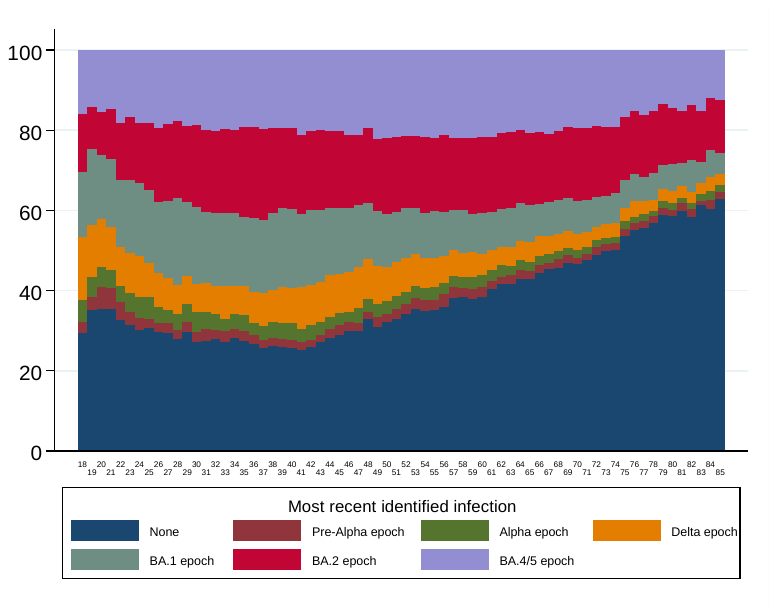

**Supplementary Figure 6. Percentages of participants’ most recent previous infection by age using last visit from 1 August 2022 to 12 September 2022.**

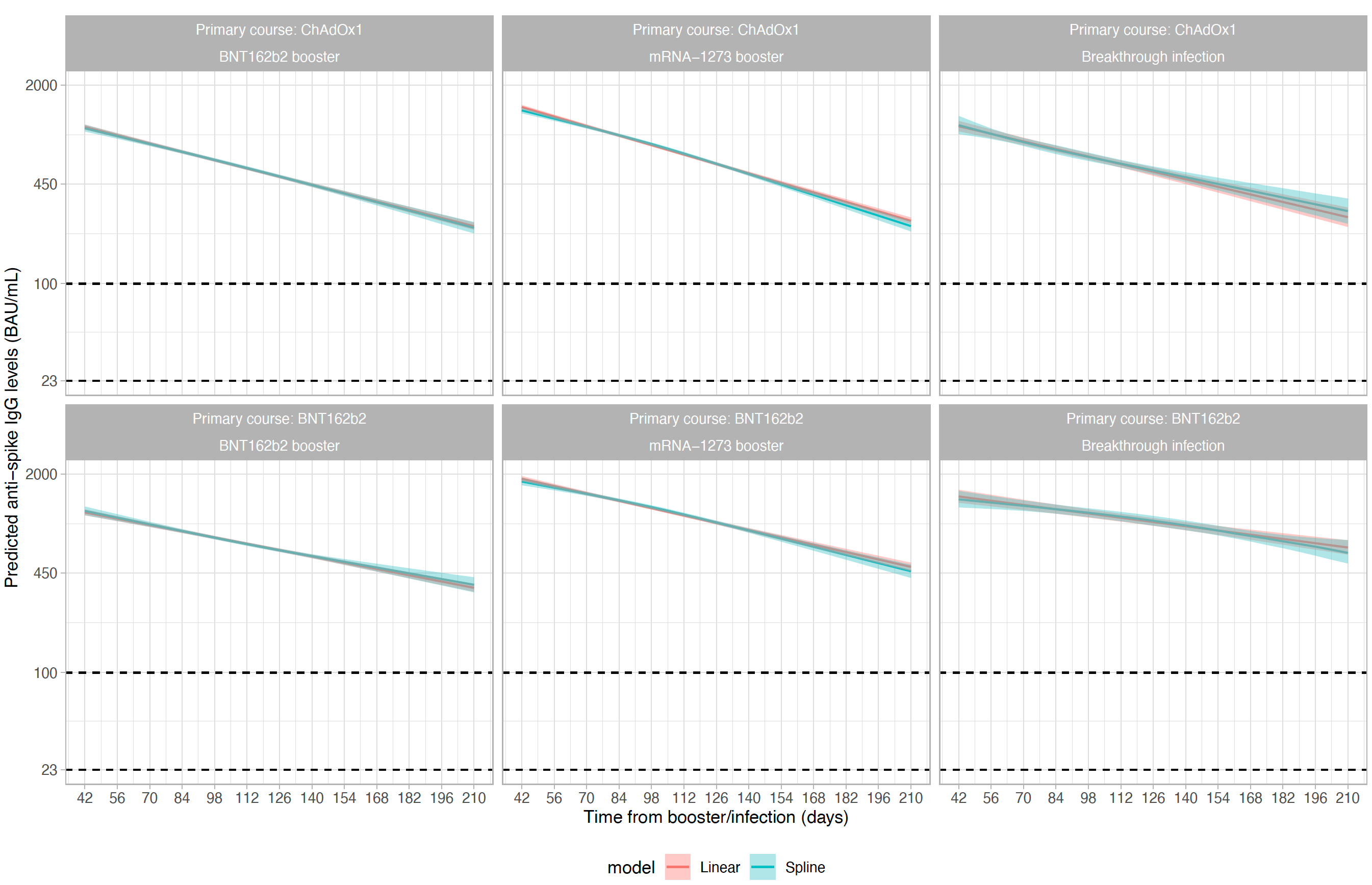

**Supplementary Figure 7. Comparison of linear exponential model with spline-based model in examining non-linearity of antibody decline.** The estimated trajectory from the spline model (with 3 knots placed at 10th, 50th, and 90th of observed time points) is similar with the linear exponential model for all six groups, indicating that there was no evidence of antibody decline flattening.

|  |  | Negative visits | Positive visits | % | Combined negative visits | Combined positive visits | % |
| --- | --- | --- | --- | --- | --- | --- | --- |
| All infection | Vaccinated without infection | 100,973 | 5,680 | 5.3 |  |  |  |
|  | Vaccinated with a most recent Pre-Alpha infection | 5,730 | 215 | 3.6 | 12,536 | 473 | 3.6 |
|  | Vaccinated with a most recent Alpha infection | 6,806 | 258 | 3.7 |  |  |  |
|  | Vaccinated with a most recent Delta infection | 15,280 | 465 | 3.0 | 41,487 | 994 | 2.3 |
|  | Vaccinated with a most recent Omicron BA.1 infection | 26,207 | 529 | 2.0 |  |  |  |
|  | Vaccinated with a most recent Omicron BA.2 infection | 38,022 | 201 | 0.5 |  |  |  |
| Ct<30 infection | Vaccinated without infection | 102,484 | 4,169 | 3.9 |  |  |  |
|  | Vaccinated with a most recent Pre-Alpha infection | 5,790 | 155 | 2.6 | 12,681 | 328 | 2.5 |
|  | Vaccinated with a most recent Alpha infection | 6,891 | 173 | 2.4 |  |  |  |
|  | Vaccinated with a most recent Delta infection | 15,430 | 315 | 2.0 | 41,811 | 670 | 1.6 |
|  | Vaccinated with a most recent Omicron BA.1 infection | 26,381 | 355 | 1.3 |  |  |  |
|  | Vaccinated with a most recent Omicron BA.2 infection | 38,098 | 125 | 0.3 |  |  |  |
| Symptomatic infection | Vaccinated without infection | 104,780 | 1,873 | 1.8 |  |  |  |
|  | Vaccinated with a most recent Pre-Alpha infection | 5,876 | 69 | 1.2 | 12,864 | 145 | 1.1 |
|  | Vaccinated with a most recent Alpha infection | 6,988 | 76 | 1.1 |  |  |  |
|  | Vaccinated with a most recent Delta infection | 15,620 | 125 | 0.8 | 42,224 | 257 | 0.6 |
|  | Vaccinated with a most recent Omicron BA.1 infection | 26,604 | 132 | 0.5 |  |  |  |
|  | Vaccinated with a most recent Omicron BA.2 infection | 38,161 | 62 | 0.2 |  |  |  |

**Supplementary Table 1. Proportion of study visits with positive PCR results in the correlates of protection analysis.** Three outcomes are examined: all infection in the Omicron BA.4/5 epoch; infections with a moderate to high viral load (Ct<30) in the Omicron BA.4/5 epoch; infections with a self-reported symptom in the Omicron BA.4/5 epoch. In the main model, pre-Alpha and Alpha, and Delta and Omicron BA.1 infections were combined because the estimates were similar.

|  | ChAdOx1-BNT162b2 (N=64940) | ChAdOx1-mRNA-1273 (N=21960) | | ChAdOx1-Infection (N=10830) | | BNT162b2-BNT162b2 (N=44197) | | BNT162b2-mRNA-1273 (N=7248) | | BNT162b2-Infection (N=4974) | | Total (N=154149) | | p value |
| --- | --- | --- | --- | --- | --- | --- | --- | --- | --- | --- | --- | --- | --- | --- |
| Months from second vaccination to booster/infection | |  | |  | |  | |  | |  | |  | |  |
| Median | 6 | 6 | | 5 | | 6 | | 6 | | 5 | | 6 | |  |
| Q1, Q3 | 6, 7 | 6, 6 | | 4, 6 | | 6, 7 | | 5, 7 | | 4, 6 | | 6, 7 | |  |
| Age (years) |  |  | |  | |  | |  | |  | |  | | < 0.001 |
| Median | 63 | 56 | | 50 | | 66 | | 41 | | 39 | | 60 | |  |
| Q1, Q3 | 53, 71 | 49, 62 | | 43, 59 | | 52, 74 | | 34, 60 | | 32, 56 | | 50, 69 | |  |
| Sex |  |  | |  | |  | |  | |  | |  | | < 0.001 |
| Female | 34760 (53.5%) | 11571 (52.7%) | | 5757 (53.2%) | | 25356 (57.4%) | | 3843 (53.0%) | | 2793 (56.2%) | | 84080 (54.5%) | |  |
| Male | 30180 (46.5%) | 10389 (47.3%) | | 5073 (46.8%) | | 18841 (42.6%) | | 3405 (47.0%) | | 2181 (43.8%) | | 70069 (45.5%) | |  |
| Ethnicity |  |  | |  | |  | |  | |  | |  | | < 0.001 |
| Non-white | 2363 (3.6%) | 844 (3.8%) | | 504 (4.7%) | | 2043 (4.6%) | | 351 (4.8%) | | 332 (6.7%) | | 6437 (4.2%) | |  |
| White | 62577 (96.4%) | 21116 (96.2%) | | 10326 (95.3%) | | 42154 (95.4%) | | 6897 (95.2%) | | 4642 (93.3%) | | 147712 (95.8%) | |  |
| Report having a long-term health condition | | |  | |  | |  | |  | |  | |  | |
| No | 45834 (70.6%) | 17428 (79.4%) | | 8684 (80.2%) | | 29429 (66.6%) | | 5635 (77.7%) | | 3964 (79.7%) | | 110974 (72.0%) | |  |
| Yes | 19106 (29.4%) | 4532 (20.6%) | | 2146 (19.8%) | | 14768 (33.4%) | | 1613 (22.3%) | | 1010 (20.3%) | | 43175 (28.0%) | |  |
| Healthcare worker |  |  | |  | |  | |  | |  | |  | | < 0.001 |
| No | 64184 (98.8%) | 21817 (99.3%) | | 10701 (98.8%) | | 41365 (93.6%) | | 7181 (99.1%) | | 4703 (94.6%) | | 149951 (97.3%) | |  |
| Yes | 756 (1.2%) | 143 (0.7%) | | 129 (1.2%) | | 2832 (6.4%) | | 67 (0.9%) | | 271 (5.4%) | | 4198 (2.7%) | |  |
| Infection type |  |  | |  | |  | |  | |  | | **Total (N=15804)** | |  |
| Delta |  |  | | 9696 (89.5%) | |  | |  | | 3194 (64.3%) | | 12890 (81.6%) | | < 0.001 |
| Omicron, BA.1 | |  | | 1119 (10.3%) | |  | |  | | 1596 (32.1%) | | 2715 (17.2%) | |  |
| Omicron, BA.2 | |  | | 13 (0.1%) | |  | |  | | 180 (3.6%) | | 193 (1.2%) | |  |
| Other |  |  | | 2 (0.0%) | |  | |  | | 0 (0.0%) | | 2 (0.0%) | |  |
| N-Miss |  |  | | 0 | |  | |  | | 4 | | 4 | |  |
| Ct value |  |  | |  | |  | |  | |  | |  | | < 0.001 |
| Median |  |  | | 20 | |  | |  | | 21 | | 20 | |  |
| Q1, Q3 |  |  | | 16, 26 | |  | |  | | 17, 26 | | 17, 26 | |  |
| Symptom |  |  | |  | |  | |  | |  | |  | | < 0.001 |
| No |  |  | | 2594 (24.1%) | |  | |  | | 1451 (29.4%) | | 4045 (25.8%) | |  |
| Yes, classic symptoms | |  | | 4444 (41.3%) | |  | |  | | 1782 (36.1%) | | 6226 (39.6%) | |  |
| Yes, other symptoms | |  | | 3735 (34.7%) | |  | |  | | 1701 (34.5%) | | 5436 (34.6%) | |  |
| N-Miss |  |  | | 57 | |  | |  | | 40 | | 97 | |  |

**Supplementary Table 2. Characteristics of all participants included in the antibody trajectory analyses.** Participants were divided into six groups based on the primary vaccine course (ChAdOx1 or BNT162b2) and the third/booster vaccination (BNT162b2 or mRNA-1273) or infection.

|  | ChAdOx1-BNT162b2 (N=41152) | ChAdOx1-mRNA-1273 (N=14748) | | ChAdOx1-Infection (N=4123) | | BNT162b2-BNT162b2 (N=24749) | | BNT162b2-mRNA-1273 (N=4403) | | BNT162b2-Infection (N=1834) | | Total (N=91009) | | p value |
| --- | --- | --- | --- | --- | --- | --- | --- | --- | --- | --- | --- | --- | --- | --- |
| Months from second vaccination to booster/infection | |  | |  | |  | |  | |  | |  | |  |
| Median | 6 | 6 | | 4 | | 6 | | 6 | | 4 | | 6 | |  |
| Q1, Q3 | 6, 7 | 6, 6 | | 3, 4 | | 6, 7 | | 5, 7 | | 3, 5 | | 6, 7 | |  |
| Age (years) |  |  | |  | |  | |  | |  | |  | | < 0.001 |
| Median | 63 | 57 | | 50 | | 66 | | 46 | | 39 | | 61 | |  |
| Q1, Q3 | 55, 71 | 51, 62 | | 43, 59 | | 55, 74 | | 35, 63 | | 33, 54 | | 52, 70 | |  |
| Sex |  |  | |  | |  | |  | |  | |  | | < 0.001 |
| Female | 22213 (54.0%) | 7838 (53.1%) | | 2254 (54.7%) | | 14145 (57.2%) | | 2345 (53.3%) | | 1036 (56.5%) | | 49831 (54.8%) | |  |
| Male | 18939 (46.0%) | 6910 (46.9%) | | 1869 (45.3%) | | 10604 (42.8%) | | 2058 (46.7%) | | 798 (43.5%) | | 41178 (45.2%) | |  |
| Ethnicity |  |  | |  | |  | |  | |  | |  | | < 0.001 |
| Non-white | 1249 (3.0%) | 507 (3.4%) | | 193 (4.7%) | | 958 (3.9%) | | 179 (4.1%) | | 121 (6.6%) | | 3207 (3.5%) | |  |
| White | 39903 (97.0%) | 14241 (96.6%) | | 3930 (95.3%) | | 23791 (96.1%) | | 4224 (95.9%) | | 1713 (93.4%) | | 87802 (96.5%) | |  |
| Report having a long-term health condition | | |  | |  | |  | |  | |  | |  | |
| No | 29072 (70.6%) | 11662 (79.1%) | | 3271 (79.3%) | | 16452 (66.5%) | | 3360 (76.3%) | | 1483 (80.9%) | | 65300 (71.8%) | |  |
| Yes | 12080 (29.4%) | 3086 (20.9%) | | 852 (20.7%) | | 8297 (33.5%) | | 1043 (23.7%) | | 351 (19.1%) | | 25709 (28.2%) | |  |
| Healthcare worker |  |  | |  | |  | |  | |  | |  | | < 0.001 |
| No | 40725 (99.0%) | 14655 (99.4%) | | 4056 (98.4%) | | 23379 (94.5%) | | 4362 (99.1%) | | 1704 (92.9%) | | 88881 (97.7%) | |  |
| Yes | 427 (1.0%) | 93 (0.6%) | | 67 (1.6%) | | 1370 (5.5%) | | 41 (0.9%) | | 130 (7.1%) | | 2128 (2.3%) | |  |
| Infection type |  |  | |  | |  | |  | |  | | **Total (N=5957)** | |  |
| Delta |  |  | | 3869 (93.8%) | |  | |  | | 1183 (64.5%) | | 5052 (84.8%) | | < 0.001 |
| Omicron, BA.1 | |  | | 242 (5.9%) | |  | |  | | 544 (29.7%) | | 786 (13.2%) | |  |
| Omicron, BA.2 | |  | | 10 (0.2%) | |  | |  | | 106 (5.8%) | | 116 (1.9%) | |  |
| Other |  |  | | 2 (0.0%) | |  | |  | | 0 (0.0%) | | 2 (0.0%) | |  |
| N-Miss |  |  | | 0 | |  | |  | | 1 | | 1 | |  |
| Ct value |  |  | |  | |  | |  | |  | |  | | < 0.001 |
| Median |  |  | | 20 | |  | |  | | 21 | | 20 | |  |
| Q1, Q3 |  |  | | 16, 26 | |  | |  | | 17, 26 | | 17, 26 | |  |
| Symptom |  |  | |  | |  | |  | |  | |  | | < 0.001 |
| No |  |  | | 942 (22.9%) | |  | |  | | 512 (28.1%) | | 1454 (24.5%) | |  |
| Yes, classic symptoms | |  | | 1854 (45.2%) | |  | |  | | 727 (39.9%) | | 2581 (43.5%) | |  |
| Yes, other symptoms | |  | | 1310 (31.9%) | |  | |  | | 583 (32.0%) | | 1893 (31.9%) | |  |
| N-Miss |  |  | | 17 | |  | |  | | 12 | | 29 | |  |

**Supplementary Table 3. Characteristics of participants included in the antibody decline analyses post third/booster vaccination or infection.** Participants were divided into six groups based on the primary vaccine course (ChAdOx1 or BNT162b2) and the third/booster vaccination (BNT162b2 or mRNA-1273) or infection. Characteristics in this subgroup are similar to that of the overall population (**Supplementary Table 2**).

| Age  (years) | Primary vaccine | Booster/Infection | Median antibody levels at 42 days (95% Crl) (BAU/mL) | Median half-life (95%Crl) (days) | Median days from booster/infection to reaching antibody associated with 67% protection (IQR) |
| --- | --- | --- | --- | --- | --- |
| 30 | BNT162b2 | mRNA-1273 | 2144 (1905-2418) | 100 (84-125) | 124 (116-133) |
|  |  | Infection | 1968 (1683-2308) | 120 (93-163) | 315 (265-411) |
| 40 | ChAdOX1 | BNT162b2 | 1441 (1300-1530) | 84 (78-90) | 57 (44-63) |
|  |  | mRNA-1273 | 1940 (1820-2064) | 69 (64-74) | 83 (78-88) |
|  |  | Infection | 1351 (1156-1583) | 73 (60-95) | 171 (155-197) |
|  | BNT162b2 | BNT162b2 | 1558 (1482-1638) | 95 (88-102) | 69 (63-74) |
|  |  | mRNA-1273 | 1940 (1774-2121) | 101 (88-117) | 103 (95-109) |
|  |  | Infection | 1630 (1387-1913) | 134 (100-207) | 308 (258-404) |
| 55 | ChAdOX1 | BNT162b2 | 1497 (1345-1572) | 78 (72-86) | 50 (0-53) |
|  |  | mRNA-1273 | 2090 (1995-2190) | 63 (61-66) | 77 (74-81) |
|  |  | Infection | 1473 (1250-1741) | 83 (63-118) | 181 (158-221) |
|  | BNT162b2 | BNT162b2 | 1402 (1326-1482) | 98 (91-106) | 0 (0-48) |
|  |  | mRNA-1273 | 2015 (1846-2200) | 93 (82-107) | 89 (81-96) |
|  |  | Infection | 1336 (1112-1610) | 141 (91-285) | 261 (196-461) |
| 70 | ChAdOX1 | BNT162b2 | 1199 (1104-1263) | 96 (88-107) | 0 (0-0) |
|  |  | mRNA-1273 | 1952 (1789-2127) | 67 (62-73) | 0 (0-50) |
|  |  | Infection | 1549 (1244-1952) | 70 (45-145) | 141 (114-217) |
|  | BNT162b2 | BNT162b2 | 1250 (1180-1326) | 110 (101-121) | 0 (0-0) |
|  |  | mRNA-1273 | 2430 (2093-2817) | 81 (69-98) | 68 (53-80) |
|  |  | Infection | 1170 (905-1518) | 117 (60-NE) | 180 (113-NE) |

**Supplementary Table 4. Posterior predicted anti-spike IgG levels at 42 days from booster/infection (BAU/mL), half-lives after booster/infection (days), and time from booster/infection to reaching antibody levels associated with 67% protection.** Results were separated by age (30, 40, 55, 70y), primary vaccine course (ChAdOx1 or BNT162b2), and booster/third vaccination or infection (BNT162b2, mRNA-1273, infection). Results were estimated at the reference category (female, white ethnicity, 6 months between second vaccination and booster/infection, not reporting a long-term health condition, not working in healthcare). Comparisons are also plotted in **Figure 3**. NE: Not estimable, due to the antibody levels not declining in the posterior median or upper credible interval.

|  | Median duration (days) between second dose and booster/infection (IQR)[range] | Selected duration (days) between second dose and booster/infection (10^th^+90^th^ percentile) | Median age (years) (IQR)[range] after using selected duration range | Age spline (years) (10^th^,50^th^,90^th^ percentile) | Time cut-off after the booster/infection (days) (90^th^ percentile, cut further for some groups 90^th^ percentile) | Number of participants  (Total/Decline model) |
| --- | --- | --- | --- | --- | --- | --- |
| ChAdOx1-BNT162b2 | 189 (183-197) [40-439] | 170-210 | 63 (53-70) [17-85] | 45, 60, 75 | 210 | 64940/41165 |
| ChAdOx1-mRNA-1273 | 187 (182-197) [72-437] | 170-210 | 56 (49-62) [17-85] | 45, 55, 65 | 210 | 21960/14748 |
| ChAdOx1-infection | 143 (98-176) [1-440] | 60-200 | 50 (43-59) [17-85] | 40, 50, 70 | 160 for age<50, 120 for age >=50 and age<70, 90 for age>=70 | 10830/4123 |
| BNT162b2-BNT162b2 | 191 (184-202) [46-475] | 150-220 | 66 (52-74) [16-85] | 40, 65, 80 | 210 | 44197/24767 |
| BNT162b2-mRNA-1273 | 180 (147-201) [67-473] | 130-220 | 41 (34-60) [17-85] | 30, 40, 70 | 210 | 7248/4408 |
| BNT162b2-infection | 140 (108-177) [1-483] | 70-220 | 39 (32-55) [16-85] | 30, 40, 70 | 180 for age<50, 120 for age >=50 and age<70, 80 for age>=70 | 4974/1834 |

**Supplementary Table 5. Settings for each Bayesian linear mixed model.** For each group, two separate models are fitted: 1) piecewise model on antibody decline after the second vaccination and subsequent increase after third/booster vaccination or infection; 2) antibody decline 42 days after the third/booster vaccination or infection. Only participants with a time between second vaccination to booster/infection from the 10^th^ to 90^th^ percentile were included in the models to avoid outlier influence. Age splines were included in the models to account for non-linearity in the association between age and antibody response. Time thresholds were applied to avoid outlier influence, using the 90^th^ percentile of the observed t>0 time points. For the older age groups in the infection models, the overall 90^th^ threshold did not exclude outliers, so we further excluded measurements outside the 90^th^ percentile in these age groups.

| Model term | Priors |
| --- | --- |
| Intercept | normal (10, 2) |
| Slope | normal (0, 0.5) |
| Coefficient for change in intercept (age) | normal (0, 1) |
| Coefficient for change in slope (age) | normal (0, 0.1) |
| Coefficient for change in intercept (duration) | normal (0, 1) |
| Coefficient for change in slope (duration) | normal (0, 0.1) |
| Coefficient for change in intercept (sex) | normal (0, 1) |
| Coefficient for change in slope (sex) | normal (0, 0.1) |
| Coefficient for change in intercept (ethnicity) | normal (0, 1) |
| Coefficient for change in slope (ethnicity) | normal (0, 0.1) |
| Coefficient for change in intercept (long-term health condition) | normal (0, 1) |
| Coefficient for change in slope (long-term health condition) | normal (0, 0.1) |
| Coefficient for change in intercept (healthcare worker) | normal (0, 1) |
| Coefficient for change in slope (healthcare worker) | normal (0, 0.1) |
| Random effect SD: intercept | normal (0, 1) |
| Random effect SD: slope | normal (0, 0.1) |
| Random effect intercept & slope covariance | lkj_corr_cholesky(1) |

**Supplementary Table 6. Priors used in the Bayesian linear mixed interval-censored models on estimating antibody decline 42 days after third/booster vaccination or infection.**

|  | | ChAdOx1 primary course | | | | | | | | | | | BNT162b2 primary course | | | | | | | | | | |
| --- | --- | --- | --- | --- | --- | --- | --- | --- | --- | --- | --- | --- | --- | --- | --- | --- | --- | --- | --- | --- | --- | --- | --- |
| BNT162b2 booster | | | | **Estimate** | **l-95% CI** | | | **u-95% CI** | | | **Rhat** | | | **Estimate** | | | **l-95% CI** | | | **u-95% CI** | | | **Rhat** |
| Population-level effects | | |  | | | |  | | |  | | | | |  | | |  | | |  | | |
| Intercept | | | | 10.1833 | 10.0771 | | | 10.2395 | | | 1.00 | | | 10.3093 | | | 10.2538 | | | 10.3647 | | | 1.01 |
| T3 | | | | -0.0848 | -0.0899 | | | -0.0740 | | | 1.02 | | | -0.0741 | | | -0.0793 | | | -0.0690 | | | 1.01 |
| age1 | | | | -0.2053 | -0.2760 | | | -0.1368 | | | 1.00 | | | -0.4625 | | | -0.5886 | | | -0.3377 | | | 1.00 |
| age2 | | | | -0.4118 | -0.4849 | | | -0.3213 | | | 1.00 | | | -0.2543 | | | -0.3820 | | | -0.1259 | | | 1.00 |
| Male1 | | | | -0.3397 | -0.3863 | | | -0.2692 | | | 1.00 | | | 0.0048 | | | -0.0551 | | | 0.0639 | | | 1.01 |
| ethnicity1 | | | | -0.0139 | -0.1193 | | | 0.0964 | | | 1.01 | | | 0.1252 | | | -0.0204 | | | 0.2729 | | | 1.00 |
| lthc1 | | | | -0.0454 | -0.0963 | | | 0.0024 | | | 1.01 | | | -0.0357 | | | -0.0977 | | | 0.0272 | | | 1.00 |
| hcw1 | | | | -0.0984 | -0.3189 | | | 0.1162 | | | 1.00 | | | -0.4434 | | | -0.6552 | | | -0.2210 | | | 1.03 |
| dur30 | | | | 0.2562 | 0.1458 | | | 0.3345 | | | 1.01 | | | 0.1283 | | | 0.0651 | | | 0.1908 | | | 1.00 |
| T3:age1 | | | | 0.0066 | -0.0001 | | | 0.0131 | | | 1.01 | | | 0.0165 | | | 0.0054 | | | 0.0279 | | | 1.00 |
| T3:age2 | | | | 0.0269 | 0.0192 | | | 0.0330 | | | 1.00 | | | 0.0174 | | | 0.0069 | | | 0.0280 | | | 1.00 |
| T3:Male1 | | | | 0.0022 | -0.0052 | | | 0.0065 | | | 1.00 | | | -0.0071 | | | -0.0124 | | | -0.0019 | | | 1.01 |
| T3:ethnicity1 | | | | 0.0066 | -0.0035 | | | 0.0171 | | | 1.01 | | | 0.0095 | | | -0.0036 | | | 0.0228 | | | 1.00 |
| T3:lthc1 | | | | -0.0025 | -0.0067 | | | 0.0028 | | | 1.00 | | | -0.0022 | | | -0.0077 | | | 0.0033 | | | 1.00 |
| T3:hcw1 | | | | 0.0034 | -0.0163 | | | 0.0216 | | | 1.01 | | | 0.0290 | | | 0.0112 | | | 0.0461 | | | 1.03 |
| T3:dur30 | | | | -0.0085 | -0.0155 | | | 0.0033 | | | 1.01 | | | 0.0030 | | | -0.0027 | | | 0.0087 | | | 1.00 |
| Group-level effects |  | | | | |  | | |  | | |  | | | |  | | |  | | |  | |
| sd(Intercept) | | | | 0.9699 | 0.0000 | | | 1.1859 | | | 1.01 | | | 1.0346 | | | 0.9935 | | | 1.0760 | | | 1.01 |
| sd(T3) | | | | 0.0767 | 0.0679 | | | 0.1092 | | | 1.01 | | | 0.0782 | | | 0.0742 | | | 0.0823 | | | 1.02 |
| cor(Intercept,T3) | | | | -0.3021 | -0.4485 | | | -0.2519 | | | 1.02 | | | -0.3572 | | | -0.4079 | | | -0.3039 | | | 1.01 |
| sigma | | | | 0.5443 | 0.5039 | | | 0.7175 | | | 1.01 | | | 0.5205 | | | 0.5103 | | | 0.5307 | | | 1.01 |
| mRNA-1273 booster | | | | **Estimate** | **l-95% CI** | | | **u-95% CI** | | | **Rhat** | | | **Estimate** | | | **l-95% CI** | | | **u-95% CI** | | | **Rhat** |
| Population-level effects | | |  | | | |  | | |  | | | | |  | | |  | | |  | | |
| Intercept | | | | 10.5452 | 10.4897 | | | 10.6000 | | | 1.00 | | | 10.7881 | | | 10.6642 | | | 10.9161 | | | 1.00 |
| T3 | | | | -0.1055 | -0.1106 | | | -0.1005 | | | 1.00 | | | -0.0696 | | | -0.0830 | | | -0.0561 | | | 1.00 |
| age1 | | | | 0.0611 | -0.0169 | | | 0.1383 | | | 1.00 | | | -0.2368 | | | -0.5039 | | | 0.0222 | | | 1.00 |
| age2 | | | | -0.0279 | -0.0898 | | | 0.0355 | | | 1.01 | | | 0.2146 | | | 0.0129 | | | 0.4160 | | | 1.00 |
| Male1 | | | | -0.2279 | -0.2794 | | | -0.1739 | | | 1.00 | | | 0.2616 | | | 0.1636 | | | 0.3587 | | | 1.00 |
| ethnicity1 | | | | -0.0165 | -0.1629 | | | 0.1272 | | | 1.00 | | | 0.1780 | | | -0.0658 | | | 0.4158 | | | 1.00 |
| lthc1 | | | | 0.0097 | -0.0568 | | | 0.0758 | | | 1.00 | | | 0.0375 | | | -0.0866 | | | 0.1630 | | | 1.00 |
| hcw1 | | | | -0.1667 | -0.4833 | | | 0.1602 | | | 1.00 | | | 0.3072 | | | -0.2513 | | | 0.8681 | | | 1.00 |
| dur30 | | | | 0.0629 | -0.0324 | | | 0.1584 | | | 1.00 | | | 0.0344 | | | -0.0499 | | | 0.1184 | | | 1.00 |
| T3:age1 | | | | -0.0088 | -0.0160 | | | -0.0015 | | | 1.00 | | | -0.0077 | | | -0.0356 | | | 0.0208 | | | 1.00 |
| T3:age2 | | | | 0.0017 | -0.0040 | | | 0.0072 | | | 1.00 | | | -0.0176 | | | -0.0371 | | | 0.0021 | | | 1.00 |
| T3:Male1 | | | | 0.0114 | 0.0065 | | | 0.0162 | | | 1.00 | | | -0.0043 | | | -0.0143 | | | 0.0059 | | | 1.00 |
| T3:ethnicity1 | | | | 0.0020 | -0.0113 | | | 0.0157 | | | 1.00 | | | -0.0020 | | | -0.0274 | | | 0.0235 | | | 1.00 |
| T3:lthc1 | | | | -0.0107 | -0.0169 | | | -0.0046 | | | 1.00 | | | -0.0155 | | | -0.0280 | | | -0.0033 | | | 1.00 |
| T3:hcw1 | | | | 0.0098 | -0.0200 | | | 0.0392 | | | 1.00 | | | -0.0051 | | | -0.0583 | | | 0.0471 | | | 1.00 |
| T3:dur30 | | | | 0.0097 | 0.0013 | | | 0.0182 | | | 1.00 | | | 0.0088 | | | -0.0001 | | | 0.0176 | | | 1.00 |
| Group-level effects |  | | | | |  | | |  | | |  | | | |  | | |  | | |  | |
| sd(Intercept) | | | | 1.1007 | 1.0681 | | | 1.1344 | | | 1.00 | | | 0.8530 | | | 0.7744 | | | 0.9298 | | | 1.00 |
| sd(T3) | | | | 0.0650 | 0.0610 | | | 0.0689 | | | 1.02 | | | 0.0645 | | | 0.0545 | | | 0.0735 | | | 1.00 |
| cor(Intercept,T3) | | | | -0.3677 | -0.4132 | | | -0.3171 | | | 1.01 | | | -0.2529 | | | -0.3892 | | | -0.0780 | | | 1.00 |
| sigma | | | | 0.5614 | 0.5506 | | | 0.5722 | | | 1.01 | | | 0.6416 | | | 0.6158 | | | 0.6687 | | | 1.00 |
| Breakthrough Infection | | | | **Estimate** | **l-95% CI** | | | **u-95% CI** | | | **Rhat** | | | **Estimate** | | | **l-95% CI** | | | **u-95% CI** | | | **Rhat** |
| Population-level effects | | |  | | | |  | | |  | | | | |  | | |  | | |  | | |
| Intercept | | | | 10.3998 | 10.1743 | | | 10.6282 | | | 1.00 | | | 10.9423 | | | 10.7172 | | | 11.1729 | | | 1.00 |
| T3 | | | | -0.0954 | -0.1172 | | | -0.0736 | | | 1.00 | | | -0.0600 | | | -0.0776 | | | -0.0428 | | | 1.00 |
| age1 | | | | 0.2514 | -0.0300 | | | 0.5319 | | | 1.00 | | | -0.9280 | | | -1.3377 | | | -0.5214 | | | 1.00 |
| age2 | | | | 0.1486 | -0.1290 | | | 0.4351 | | | 1.00 | | | -0.5804 | | | -0.9105 | | | -0.2501 | | | 1.00 |
| Male1 | | | | 0.0782 | -0.0972 | | | 0.2555 | | | 1.00 | | | 0.3148 | | | 0.0975 | | | 0.5318 | | | 1.00 |
| ethnicity1 | | | | 0.2343 | -0.1718 | | | 0.6499 | | | 1.00 | | | 0.0716 | | | -0.3648 | | | 0.5041 | | | 1.00 |
| lthc1 | | | | 0.0011 | -0.2130 | | | 0.2208 | | | 1.00 | | | -0.1926 | | | -0.4766 | | | 0.0908 | | | 1.00 |
| hcw1 | | | | -0.5783 | -1.1839 | | | 0.0445 | | | 1.00 | | | -0.3635 | | | -0.7891 | | | 0.0602 | | | 1.00 |
| dur30 | | | | 0.1970 | 0.1217 | | | 0.0445 | | | 1.00 | | | 0.2257 | | | 0.1392 | | | 0.3105 | | | 1.00 |
| T3:age1 | | | | 0.0155 | -0.0199 | | | 0.0511 | | | 1.00 | | | 0.0169 | | | -0.0219 | | | 0.0564 | | | 1.00 |
| T3:age2 | | | | -0.0125 | -0.0645 | | | 0.0375 | | | 1.00 | | | -0.0050 | | | -0.0620 | | | 0.0528 | | | 1.00 |
| T3:Male1 | | | | 0.0025 | -0.0180 | | | 0.0228 | | | 1.00 | | | -0.0200 | | | -0.0380 | | | -0.0021 | | | 1.00 |
| T3:ethnicity1 | | | | -0.0059 | -0.0489 | | | 0.0382 | | | 1.00 | | | 0.0165 | | | -0.0183 | | | 0.0511 | | | 1.00 |
| T3:lthc1 | | | | 0.0019 | -0.0253 | | | 0.0285 | | | 1.00 | | | -0.0013 | | | -0.0276 | | | 0.0244 | | | 1.00 |
| T3:hcw1 | | | | 0.0538 | -0.0120 | | | 0.1201 | | | 1.00 | | | 0.0456 | | | 0.0023 | | | 0.0883 | | | 1.00 |
| T3:dur30 | | | | -0.0024 | -0.0104 | | | 0.0053 | | | 1.00 | | | -0.0081 | | | -0.0151 | | | -0.0009 | | | 1.00 |
| Group-level effects |  | | | | |  | | |  | | |  | | | |  | | |  | | |  | |
| sd(Intercept) | | | | 1.7485 | 1.6314 | | | 1.8660 | | | 1.00 | | | 1.5703 | | | 1.4386 | | | 1.7069 | | | 1.00 |
| sd(T3) | | | | 0.0745 | 0.0547 | | | 0.0951 | | | 1.01 | | | 0.0665 | | | 0.0496 | | | 0.0818 | | | 1.00 |
| cor(Intercept,T3) | | | | -0.8975 | -0.9912 | | | -0.7963 | | | 1.01 | | | -0.7548 | | | -0.8420 | | | -0.6583 | | | 1.00 |
| sigma | | | | 0.9574 | 0.8992 | | | 1.0180 | | | 1.01 | | | 0.7438 | | | 0.6898 | | | 0.8023 | | | 1.00 |

**Supplementary Table 7. Model coefficients and MCMC diagnostics for the Bayesian linear mixed models estimating antibody decline 42 days after the third/booster vaccination or infection.**
